# *GBA* in Parkinson’s disease: variant detection and pathogenicity scoring matters

**DOI:** 10.1101/2023.02.03.23285410

**Authors:** Carolin Gabbert, Susen Schaake, Theresa Lüth, Christoph Much, Christine Klein, Jan O. Aasly, Matthew J. Farrer, Joanne Trinh

## Abstract

**Background:** *GBA* variants are the strongest genetic risk factor for Parkinson’s disease (PD). However, the pathogenicity of *GBA* variants concerning PD is still not fully understood. Additionally, the frequency of *GBA* variants varies widely across populations.

**Objectives:** To evaluate Oxford Nanopore sequencing as a strategy, to determine the frequency of *GBA* variants in Norwegian PD patients and controls, and to review the current literature on newly identified variants that add to pathogenicity determination.

**Methods:** We included 462 Norwegian PD patients and 367 healthy controls. We sequenced the full-length *GBA* gene on the Oxford Nanopore GridION as an 8.9 kb amplicon. Six analysis pipelines were compared using two aligners (NGMLR, Minimap2) and three variant callers (BCFtools, Clair3, Pepper-Margin-Deepvariant). Confirmation of *GBA* variants was performed by Sanger sequencing and the pathogenicity of variants was evaluated.

**Results:** We found 95.8% (115/120) true-positive *GBA* variant calls, while 4.2% (5/120) variant calls were false-positive, with the NGMLR/Minimap2-BCFtools pipeline performing best. In total, 13 rare *GBA* variants were detected: two were predicted to be (likely) pathogenic and eleven were of uncertain significance. The odds of carrying one of the two most common *GBA* variants, p.L483P or p.N409S, in PD patients were estimated to be 4.11 times the odds of carrying one of these variants in controls (OR=4.11 [1.39, 12.12]).

**Conclusions:** In conclusion, we have demonstrated that Oxford long-read Nanopore sequencing, along with the NGMLR/Minimap2-BCFtools pipeline is an effective tool to investigate *GBA* variants. Further studies on the pathogenicity of *GBA* variants are needed to assess their effect on PD.

## Introduction

Parkinson’s disease (PD) is a multifaceted and highly complex neurodegenerative disorder. Multiple genes have been implicated in PD. Variants in *GBA* (Glucocerebrosidase A) are considered a common genetic risk factor for PD^1-6^. Biallelic (homozygous or compound heterozygous) variants in *GBA* classically cause Gaucher’s disease (GD) and an increased PD risk has been observed in patients with GD and asymptomatic carriers of heterozygous variants^7-10^. Glucocerebrosidase enzymatic activity is reduced in patients with PD who carry a *GBA* heterozygous variant compared to non-carriers, and it is even lower in *GBA* homozygotes/compound heterozygotes^10^. Common *GBA* variants in PD include p.E365K (NM_000157.4, c.1093G>A), p.T408M (NM_000157.4, c.1223C>T), p.N409S (NM_000157.4, c.1226A>G), and p.L483P (NM_000157.4, c.1448T>C). However, the classification of the pathogenicity of *GBA* variants and their effect on PD is still ongoing. For this reason, the reported frequencies of *GBA* variants across studies are rather inconsistent, with frequencies ranging from 1.8% up to 47% depending on the ethnicity of the samples and the *GBA* variants investigated^2,5,11^. In a previous study on the Norwegian population, 311 patients with PD were included and screened for the two most common *GBA* variants (i.e., p.N409S and p.L483P)^12^. Seven patients (2.3%) that carried a heterozygous *GBA* variant were found: four of the patients had a p.N409S (1.3%) and three had a p.L483P (1.0%) substitution.

Another challenge that arises when sequencing the *GBA* gene is the nearby pseudogene *GBAP1. GBAP1* shares 96% exonic sequence homology with the *GBA* coding region with the highest homology between exons 8 and 11. In this region, most pathogenic variants have been reported, usually resulting from recombination events, e.g. gene conversion, fusion, or duplication^13^. This complex regional genomic structure complicates PCR and DNA sequencing. To avoid the pseudogene, one method to analyze *GBA* is by long-read sequencing^14^. This technology provides full-length *GBA* sequencing to detect exonic and intronic variants and recombinant alleles in combination with phase information, at high multiplex capacity^15,16^.

Herein, we have comprehensively characterized *GBA* in a sample from the Norwegian population by 1) employing and evaluating Oxford Nanopore sequencing as a strategy, 2) determining the frequency of variants within the *GBA* gene in patients with PD and healthy controls by providing an update on previous reports^12,17^, and 3) reviewing current literature on newly identified variants that add to pathogenicity determination.

## Methods

### Demographics

A sample of 462 Norwegian patients with PD was included in this study (Table 1). All patients were referred by general practitioners and other hospitals and have been clinically examined and observed longitudinally at the outpatient clinics of three hospitals in Central Norway. One hundred eighty (39%) of the patients were men, and 282 (61%) were women. The mean age at disease onset in the patient group was 60.3 years (SD = ± 9.7 years, range 26 to 88 years). Forty-three out of 462 patients were probands with a family history of PD. Patients with a known genetic cause of PD were not included. In addition, a group of 367 healthy Norwegian individuals (mean age 64.0 years) originating from the same geographic region and without signs of a movement disorder was included to determine the variant frequency in the general population (Table 1).

**Table 1.** Demographics of the Norwegian patients with PD and healthy controls.

| Full Cohort (n=829) | Patients with PD | Healthy Controls |
| --- | --- | --- |
| N | 462 | 367 |
| Male/Female (%) | 180/282 (39%/61%) | 159/198 (43%/54%) |
| Mean AAO (SD, range) | 60.3 (±9.7, 26-88) | NA |
| Mean Age (SD, range) | NA | 64.0 (±12.2, 30-96) |
| Family history of PD | 43 | 0 |

### Genetic analysis

#### Long-read Oxford Nanopore Sequencing

We used blood-derived genomic DNA samples from all PD patients and controls. Informed consent was obtained from all participating individuals. We enriched for *GBA* by amplifying an 8.9 kb sequence, which covered all coding exons, the introns between them, and part of the 3’ UTR region (hg38: chr1:155232501-155241415), described previously^15,16^ using the LongAmp Taq PCR Kit. Subsequently, 1.3 μg of each patient-derived PCR product was barcoded with the Native 96 Barcoding Kit (EXP-NBD196) and multiplexed. The libraries were generated with the Ligation Sequencing Kit (SQK-LSK109) for long-read Nanopore sequencing on R9.4.1 flow cells (FLO-MIN106) on a GridION.

#### Bioinformatic analyses

Data acquisition and run monitoring was carried out with MinKNOW (version v21.05.25 and later). The integrated Guppy algorithm (version v5.0.16 and later) was used for base-calling with the super-accurate base-calling model, de-multiplexing, and FAST5 and FASTQ file generation. The base-called reads were filtered with Filtlong (v.0.2.0) (https://github.com/rrwick/Filtlong) to only include the best 50% of the reads, based on Phred quality scores (q-score) in the FASTQ files, with a minimum read length of 8 kb. Afterwards, the reads were trimmed with NanoFilt^22^ (v2.8.0) and 75 bp were cropped from the front of the reads and 20 bp from the end. Subsequently, the nanopore reads were aligned against the reference sequence (hg38). We used two different aligners: NGMLR^23^ (v0.2.7) and Minimap2^24^ (v2.22). Then, the alignments were sorted and indexed with SAMtools^25^ using v1.9 for the NGMLR alignment, and v1.15 for the Minimap2 alignment. In addition, the coverage for each sample was calculated using SAMtools^25^ (v.1.15). The processed BAM files were analyzed with three different variant callers: BCFtools^25^ (v1.9), Clair3 (v0.1-r11), and the Pepper-Margin-Deepvariant pipeline^26^ (Supplementary Figure 1). Finally, the resulting VCF files containing the SNPs within *GBA* were annotated using ANNOVAR^27^ (version 2020-06-11).

**Figure 1.**
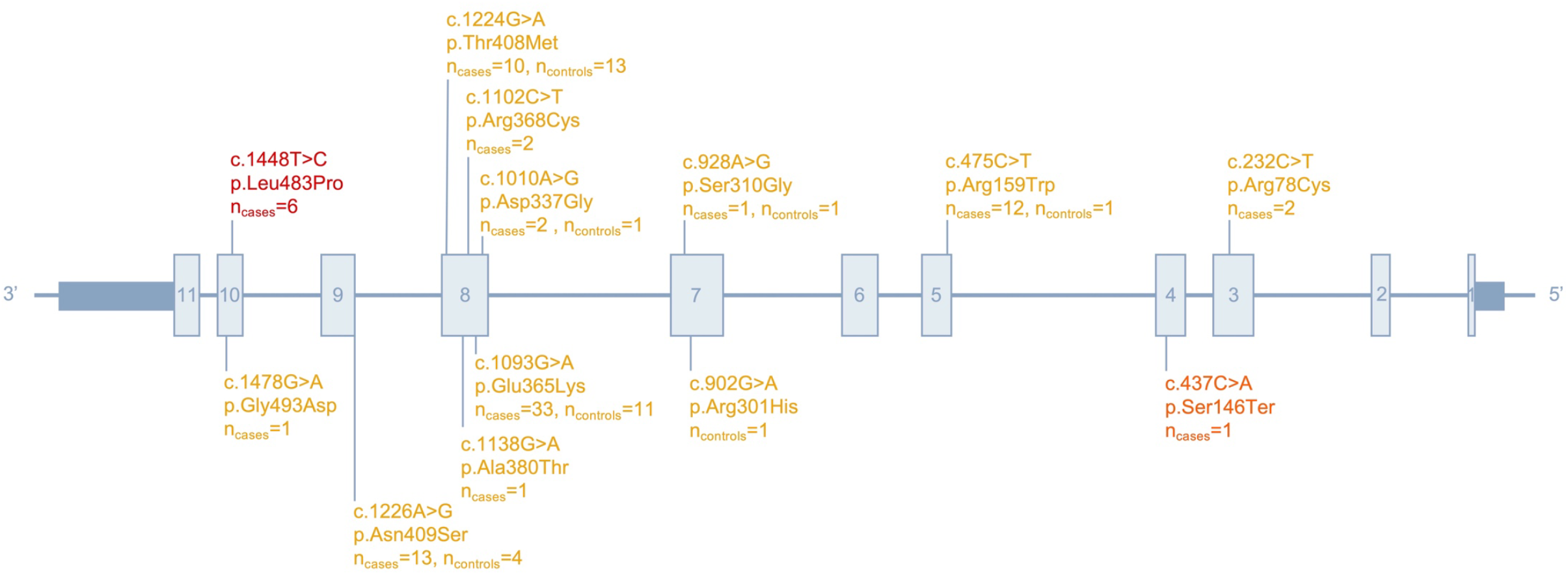
Schematic representation of the exonic (gray boxes) and intronic (gray lines) structure of the *GBA* gene highlighting “pathogenic” (red)/“likely pathogenic” (orange)/“uncertain” (yellow) variants found in our Norwegian series.

#### Sanger sequencing and structural variant detection

Sanger sequencing was performed for all individuals with a rare “pathogenic”/“likely pathogenic”/“uncertain” *GBA* variant, as previously described^28^. Individuals with a rare “pathogenic”/“likely pathogenic”/“uncertain” *GBA* variant were further examined for possible structural variants (SVs) due to the high exonic sequence homology between *GBA* and *GBAP1*. We used two sets of primers, as previously described^16^, to detect reciprocal crossovers between the gene and pseudogene resulting in a 20.6 kb deletion or a 20.6 kb duplication using the LongAmp Taq PCR Kit. The resulting PCR products were subsequently run in a 1.5% agarose gel.

#### Sensitivity assessment

To assess the performance of variant calling, we stratified the detected *GBA* variants into true-positive, false-positive, and false-negative calls for each data analysis pipeline (NGMLR+BCFtools, NGMLR+Clair3, NGMLR+Pepper-Margin-Deepvariant, Minimap2+BCFtools, Minimap2+Clair3, Minimap2+Pepper-Margin-Deepvariant). True-positive variants were defined as *GBA* variants detected with Nanopore sequencing that were validated with Sanger sequencing. False-positive variants are those identified with Nanopore sequencing but not confirmed with Sanger sequencing. False-negative variants were determined as those not called with a specified data analysis pipeline, but later validated with Sanger sequencing. Evaluation of each data analysis pipeline was based on the ratio of false-positive to false-negative calls.

### Pathogenicity scoring

*GBA* single nucleotide variants were first filtered based on GnomAD frequency <2%. Pathogenicity classification and scoring were assessed with American College of Medical Genetics and Genomics (ACMG)^29^ criteria. This included using Varsome^30^, ClinVar^31^, SIFT^32^, Polyphen2^33^, CADD^34^, and GERP++^35^. *GBA* variants were categorized as “pathogenic”, “likely pathogenic”, or of “uncertain significance” and further validated by Sanger sequencing. The p.E365K variant, which is a known risk factor for PD, was additionally categorized as of “uncertain significance”, despite the “likely benign” score from ACMG. Variants categorized as of “uncertain significance” without information from mutation predictors were not Sanger sequenced.

### Literature review

We performed a systematic literature review to summarize *GBA* variants detected in PD and the frequency across different populations (Supplementary Figure 2). We searched for literature via PubMed that was published before August 4, 2022, using the search term “*GBA*” AND “Parkinson” AND “prevalence” OR “*GBA*” AND “Parkinson” AND “frequency”, while setting the species filter to “Human” and the language filter to “English”, resulting in 94 articles. These were screened based on the title, abstract, and full text, excluding all articles not directly screening for variants in the *GBA* gene in patients with PD. Of these 94 articles, 41 articles were excluded. Reasons for exclusion were reviews or comments without new data (n=11), articles that did not perform *GBA* variant screening or examine *GBA* variant frequency in their study population (n=22), and articles that did not include patients with PD (n=10). Multiple reasons for exclusion were possible. In addition to the articles found via the search term, suitable articles that were referenced in this literature were also included in the overview.

## Results

### Long-read sequencing of *GBA*

After Nanopore sequencing of the long-range PCR products, we obtained a mean read length of 5.2 kb (SD = ± 2.1 kb) and a mean read quality Phred score of 14.2 (SD = ± 0.4) across all raw sequencing data. After length and quality filtering and read trimming, we obtained a mean read length of 9.0 kb (SD = ± 0.2 kb) and a mean read quality Phred score of 15.7 (SD = ± 0.5). For the filtered samples the mean coverage was 193.1X (SD ± 187.5X), ranging between 20.6X and 1820.8X across all samples. Nevertheless, the coverage per sample was consistent over all positions (Supplementary Figure 3).

### Bioinformatic pipeline comparison

In total, 79 rare *GBA* variants (gnomAD frequency <2%) were detected in the Nanopore sequencing analysis after filtering. Out of the 79 rare *GBA* variants, 18 variants were categorized as “pathogenic”, “likely pathogenic”, or of “uncertain significance” (Supplementary Table 1). The remaining 61 rare *GBA* variants were categorized as “benign” or “likely benign”. The number of *GBA* variants differed across all six analysis pipelines (Table 2).

**Table 2.** *GBA* variants sequenced with Oxford Nanopore and analyzed with six different pipelines using NGMLR and Minimap2 aligners and BCFtools, Clair3, and Pepper-Margin-Deepvariant callers. variants^*^ pathogenic”/“uncertain *GBA*

| Alignment | NGMLR |  |  | Minimap2 |  |  |
| --- | --- | --- | --- | --- | --- | --- |
| Variant calling | BCFtools | Clair3 | Pepper-Margin-Deepvariant | BCFtools | Clair3 | Pepper-Margin-Deepvariant |
| N of calls of the 79 rare *GBA* variants* | 433 | 426 | 454 | 433 | 429 | 481 |
| N of calls of the 18 rare “pathogenic”/“likely pathogenic”/“uncertain *GBA* variants** | 120 | 117 | 123 | 120 | 119 | 130 |
| N of Sanger validated calls | 115 | 112 | 108 | 115 | 113 | 109 |
| N of false-positive calls | 5 | 5 | 15 | 5 | 6 | 21 |
| N of false-negative calls | 0 | 3 | 7 | 0 | 2 | 6 |

\*Variants were annotated using Annovar; variants with a gnomAD frequency >2% and without information on SNP ID or amino acid change were excluded.
\*\*Variants were classified based on the pathogenicity reported by ACMG, Varsome, Clinvar, SIFT, Polphen2, CADD, and GERP++.

#### BCFtools

With BCFtools, 64 rare annotated *GBA* variants were detected in 313 samples, resulting in 433 calls in total. The calls were identical with both aligners (i.e., NGMLR and Minimap2). Of these, 15 rare variants, categorized as “pathogenic”, “likely pathogenic”, or of “uncertain significance”, were found in 111 samples (i.e., 120 calls), again independent of the aligner. After Sanger sequencing, 13 variants in 110 samples (i.e., 115 calls) were validated, indicating five false-positive calls but no false-negative calls.

#### Clair3

For the pipeline using Clair3 as a variant caller after a preceding alignment with NGMLR, 65 rare annotated *GBA* variants were detected in 308 samples, resulting in 426 calls. Of these calls, 117 calls referred to 14 rare variants, categorized as “pathogenic”, “likely pathogenic”, or of “uncertain significance”, and were detected in 108 samples. After Sanger sequencing, 112 calls in 107 samples were validated, implying that five calls were false-positive, with an additional three calls being false-negative. When Minimap2 was used as an aligner, again 65 rare annotated *GBA* variants were detected, however, here they were detected in 311 samples (i.e., 429 calls). With this pipeline, 15 rare annotated *GBA* variants, categorized as “pathogenic”, “likely pathogenic”, or of “uncertain significance”, were found in 110 samples (i.e., 119 calls). Of these, 113 calls in 108 samples could be validated with Sanger sequencing, indicating six false-positive calls and two more false-negative calls.

#### Pepper-Margin-Deepvariant

Lastly, the Pepper-Margin-Deepvariant pipeline was used for variant calling. After preceding alignment with NGMLR, 453 calls of 72 rare annotated *GBA* variants were detected in 318 samples, including 123 calls of 17 rare variants, categorized as “pathogenic”, “likely pathogenic”, or of “uncertain significance”, in 114 samples. Only 108 calls in 103 samples were validated with Sanger sequencing, leading to 15 false-positive and additional seven false-negative calls. Similarly, when Minimap2 was used for the alignment, 76 rare annotated *GBA* variants were detected in 331 samples, resulting in 481 calls. Of these, 18 rare variants, categorized as “pathogenic”, “likely pathogenic”, or of “uncertain significance”, were detected in 120 samples (i.e., 130 calls). Here, 109 calls in 104 samples were Sanger validated, indicating 21 false-positive and six false-negative calls.

### *GBA* variant frequencies in the Norwegian population

In total, 13/18 rare distinct variants within *GBA* were validated in 462 Norwegians with PD and 367 healthy controls (Figure 1, Supplementary Table 2), whereas 5/18 could not be validated. Two of the 13 rare *GBA* variants were predicted to be “pathogenic” or “likely pathogenic” (p.L483P, p.S146X) and eleven *GBA* variants were of “uncertain significance” (p.G493D, p.N409S, p.T408M, p.A380T, p.R368C, p.E365K, p.D337G, p.S310G, p.R301H, p.R159W, p.R78C). The total carrier frequency of rare *GBA* variants predicted as “pathogenic”, “likely pathogenic”, or of “uncertain significance” was 17.1% (79/462) in the PD cases and 8.4% (31/367) in the controls (OR=2.24 [1.44, 3.47]) (Table 2). The carrier frequency of known *GBA* risk variants for PD, including p.L483P, p.N409S, p.T408M, p.E365K, and p.R159W, was 15.2% in the PD cases and 7.9% in the controls (OR=2.08 [1.32, 3.29]) (Table 3). With regard to the two most common *GBA* risk variants for PD, the frequency of carrying either a p.L483P or p.N409S variant was 4.3% in patients with PD and 1.1% in healthy controls (OR=4.11 [1.39, 12.12]) (Table 3).

**Table 3.**
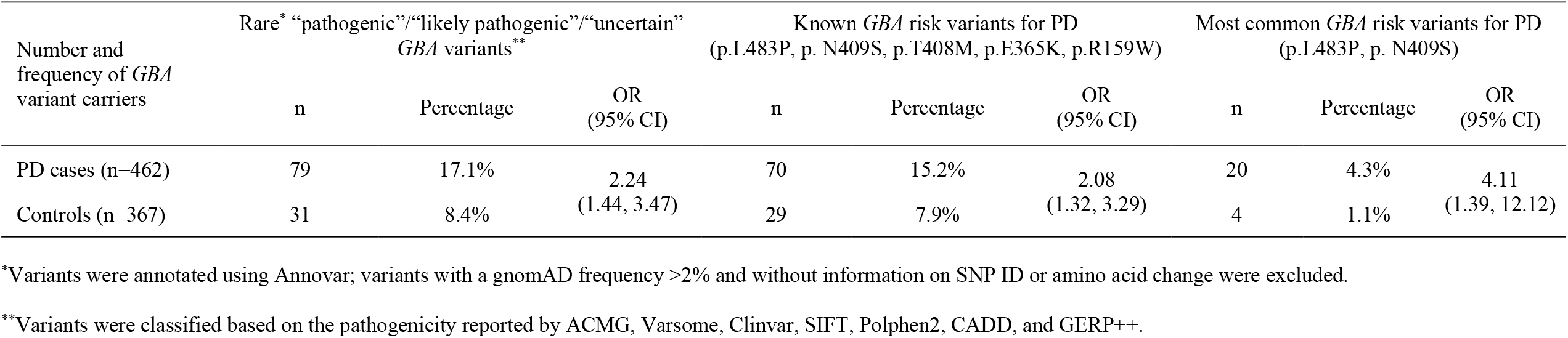
Number and frequency of patients and controls with *GBA* variants in our Norwegian series.

In addition, samples with a rare “pathogenic”, “likely pathogenic”, or “uncertain” *GBA* variant were further examined for possible structural variants (SVs). However, all samples tested negative for SVs.

### Literature review

We performed a systematic literature review to summarize *GBA* variants detected in PD and their frequencies across different populations. In total, 100 articles on *GBA* variant frequencies across populations were included in the overview (Supplementary Table 3). Most of the studies assessed the p.L483P and p.N409S variants, with p.L483P variant frequencies ranging between 0% and 8.3% for cases with PD and between 0% and 1.4% in the general population. The highest frequency in PD patients was observed in a sample from the Japanese population^18^. For p.N409S, the frequencies ranged from 0% to 26.3% in patients with PD and from 0% to 5.96% in controls, with the highest frequency reported in a sample from the Ashkenazi Jewish population^19^. In a previous study on the Norwegian population, the frequency of p.L483P was predicted to be 0.5% in patients with PD (2/442) and 1.4% in controls (6/419) (OR=0.31 [0.06, 1.56])^17^. For the p.N409S variant, the frequencies reported were 0.2% in patients with PD (1/442) which was comparable to controls (1/419) (OR=0.95 [0.06, 15.2])^17^. In addition, the study reported *GBA* p.E365K, detected in 4.3% of patients with PD (18/442) and in 6.6% of controls (29/419) (OR=0.57 [0.13, 1.04]), and p.T408M, found in 1.7% of patients with PD (7/442) and in 3.6% of controls (16/419) (OR=0.41 [0.17, 1]).

## Discussion

Variants in the *GBA* gene are known to affect PD risk, however, the frequency and pathogenicity of these variants are still under debate. The latter is further complicated as the frequencies of *GBA* variants vary by population. In our study, we used the pathogenicity scoring of ACMG, Varsome, ClinVar, SIFT, Polyphen2, CADD, and GERP++ and categorized the variants found into “pathogenic”, “likely pathogenic”, and variants of “uncertain significance”. Here we report 13 rare variants within *GBA* in Norwegian PD cases, with two of them predicted to be “pathogenic” or “likely pathogenic”, and eleven *GBA* variants of “uncertain significance”. In total, we found a “pathogenic” or “likely pathogenic” variant (e.g., p.L483P, p.S146X) in 1.5% of PD cases and in 0% of healthy controls. However, pathogenicity scoring with ACMG does not take the association with PD into account and consequently underestimates the frequency of risk variants in *GBA*. Therefore, we further investigated known *GBA* risk variants associated with PD. In our study, 15.2% of patients with PD carried a common *GBA* risk variant (i.e., p.L483P, p.N409S, p.T408M, p.E365K, and p.R159W), compared to 7.9% of controls.

In a systematic literature review, we evaluated the frequency of variants in the *GBA* gene in patients with PD and in the general population in 100 studies (Supplementary Table 3). The frequencies of *GBA* risk variants range between 0% and 26.3% in patients with PD, highlighting the importance of investigating *GBA* variants across populations. In a previous study on the Norwegian population, two known risk variants, p.L483P and p.N409S, were investigated among others^17^. The frequency of p.L483P was predicted to be 0.5% in patients with PD and 1.4% in controls, while 0.2% of the patients with PD and controls had a p.N409S variant^17^. Therefore, the odds of carrying a p.L483P or a p.N409S variant in patients with PD are lower than in healthy controls (OR=0.31 [0.06, 1.56], OR=0.95 [0.06, 15.2]). In contrast to this, we found slightly higher frequencies in our Norwegian series. The p.L483P variant was found in 1.3% of Norwegian PD cases and in 0% of controls, the p.N409S variant in 2.8% of PD patients and 1.1% of controls, leading to higher odds of carrying these variants in patients with PD compared to controls (OR=2.63 [0.85, 8.13]).

These variants are classically found in GD but are also associated with PD risk. However, some variants in *GBA* show associations with PD but do not cause GD, e.g., the *GBA* variants p.E365K and p.T408M^6^. In our Norwegian series, the p.E365K variant was found in 7.1% of cases with PD and in 3.0% of the general population (OR=2.49 [1.24, 5]). The p.T408M variant was found in 2.2% of cases with PD and in 3.5% of the general population (OR=0.6 [0.26, 1.39]). Hence, the pathogenicity of *GBA* variants p.E365K and p.T408M in PD is still unclear.

Although the pathogenicity and the pathomechanism of several *GBA* variants in patients with PD are still under discussion, gene-targeted therapy might help to treat *GBA*-PD. So far, there are several strategies for the treatment of *GBA*-PD with ongoing clinical trials. Some of the therapeutic approaches for *GBA* include substrate reduction therapy targeting glycosylceramide synthase inhibition, the inhibition of glucocerebrosidase transportation, the development of glucocerebrosidase activators, and gene therapy targeting the replacement of mutated *GBA* with WT copies of the gene^20^. In addition to variable inclusion criteria, the sequencing method and data analysis, and the chronology when it was performed, has a major influence on the detection rate and *GBA* variant frequencies reported. Through the years, DNA sequencing technologies evolved tremendously with new sequencing techniques and especially new prediction tools improving the accuracy of variant detection. Continual refinement in these tools enables more comprehensive identification of variants and highlights the need to re-evaluate known genes as time goes by. Long-read sequencing in combination with the latest data analysis tools enabled us to determine the frequency of *GBA* variants in the Norwegian population with higher precision than before. We evaluated the consensus and accuracy of six different pipelines using two different aligners (NGMLR and Minimap2), as well as three different variant callers (BCFtools, Clair3, and the Pepper-Margin-Deepvariant pipeline).

BCFtools performed best with regard to the number of true-positive, false-positive, and false-negative hits, independently from the aligner used. However, one limitation is that we could not fully evaluate pipeline sensitivity. As we only confirmed variants called by our Nanopore analysis pipelines by Sanger sequencing, we underestimate false-negative variants that were not initially called. With Oxford Nanopore technology we assessed the precision of long-read sequencing and consensus data analysis and detected 115 real *GBA* variant calls, while five variant calls were false-positives. Thus, >95% of called variants were true-positive. Nanopore long-read sequencing is an accurate tool to detect genetic variations and with further development in flow cells and sequencing kits, the accuracy of variant detection is likely to increase. Another advantage of this technology is the capacity to multiplex samples, which decreases analysis costs of the full 8.9 kb *GBA* gene to $13 USD per sample, which is lower than other DNA sequencing methods^21^. A strength of Oxford Nanopore long-range sequencing is to specifically target the *GBA* gene without sequencing the pseudogene *GBAP1*, and to detect all disease-causing variants including information on phase^15,16^.

In conclusion, we have demonstrated that Oxford Nanopore sequencing is an efficient tool for investigating *GBA* variants. The frequency of the two most common *GBA* risk variants for PD, p.L483P and p.N409S, in Norwegian patients with PD is 4.3% and higher than in the general population (1.1%). Given the importance of this gene, further functional studies on the pathogenicity of *GBA* are needed to assess their effect on PD.

## Data Availability

All data produced in the present study are available upon reasonable request to the authors.

## Acknowledgment

This project was supported by the Institute of Neurogenetics at the University of Lübeck. Grant support was received from the DFG RU ProtectMove (FOR2488). Special thanks to the families and patients who have participated in this study.

## Author contributions

Carolin Gabbert: Data analysis, Data interpretation, Writing

Susen Schaake: Acquisition of data, Data interpretation, Review and Critique

Theresa Lüth: Contributed to data analysis, Data interpretation, Review and Critique

Christoph Much: Acquisition of data, Data interpretation, Review and Critique

Christine Klein: Review and Critique

Jan Aasly: Sample acquisition, Review and Critique

Matthew J. Farrer: Review and Critique

Joanne Trinh: Conceptualization, Data analysis, Data interpretation, Writing

## Data availability

The data that support the findings of this study are available on request from the corresponding author.

## Financial Disclosures

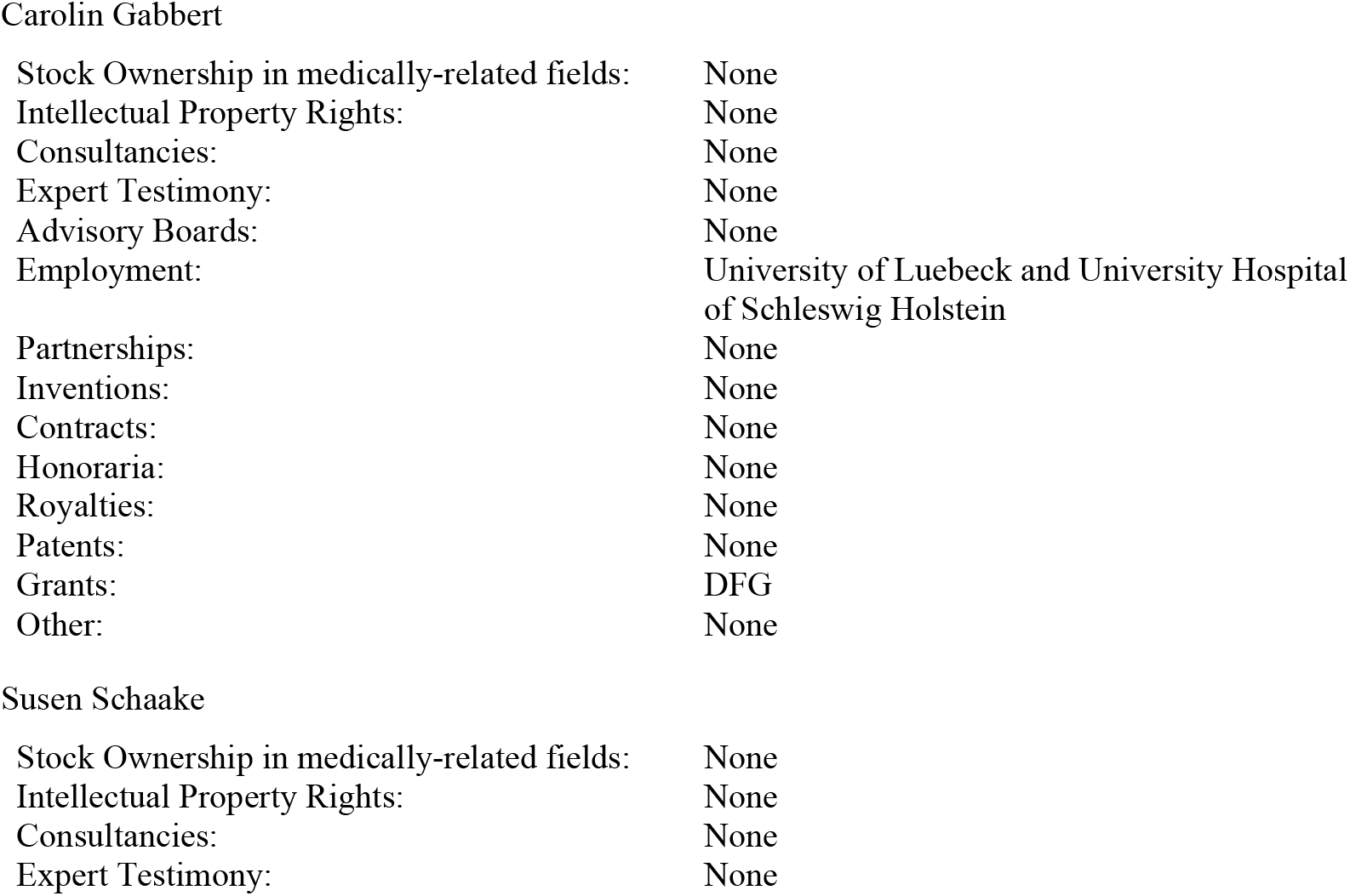
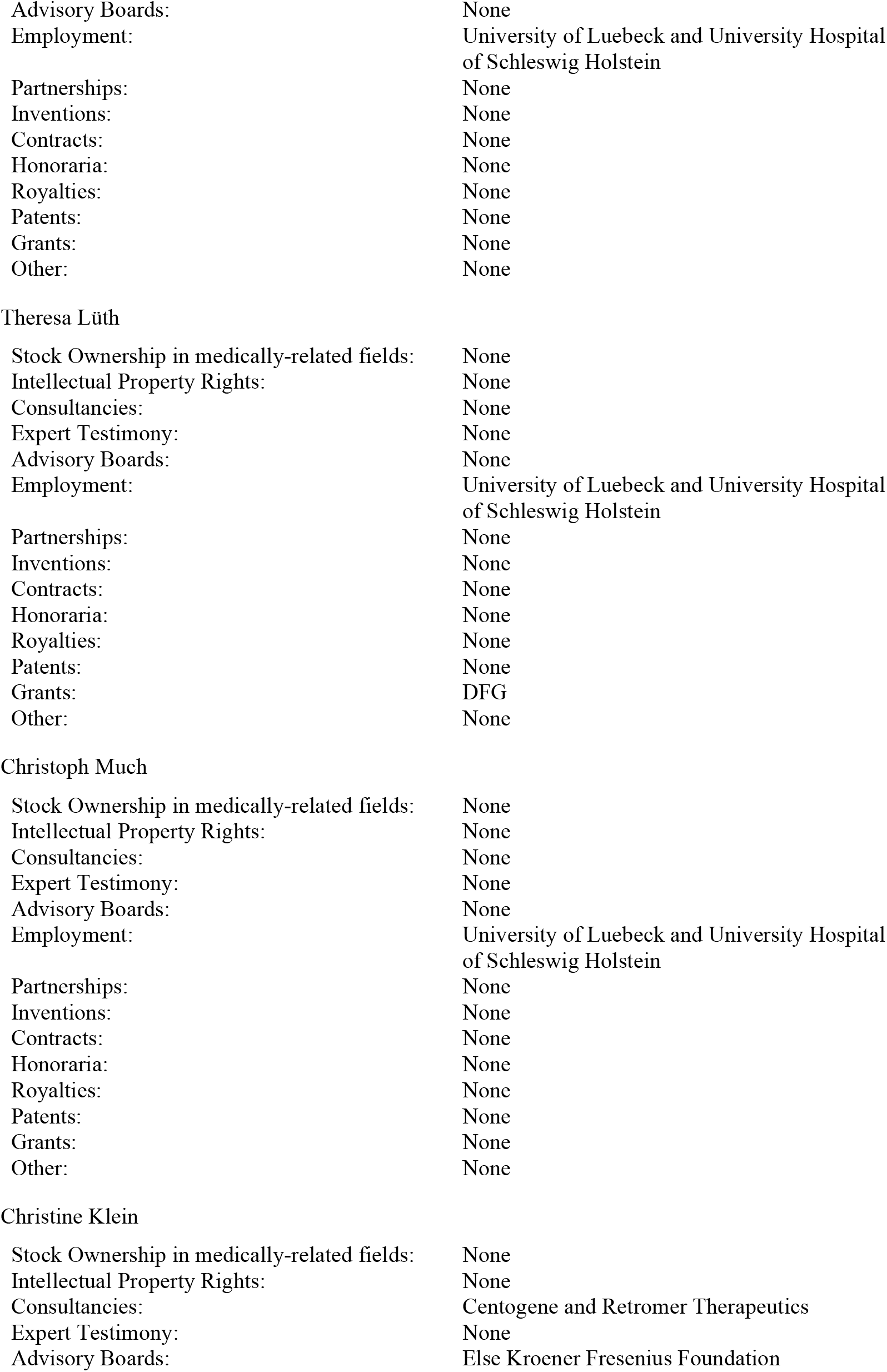
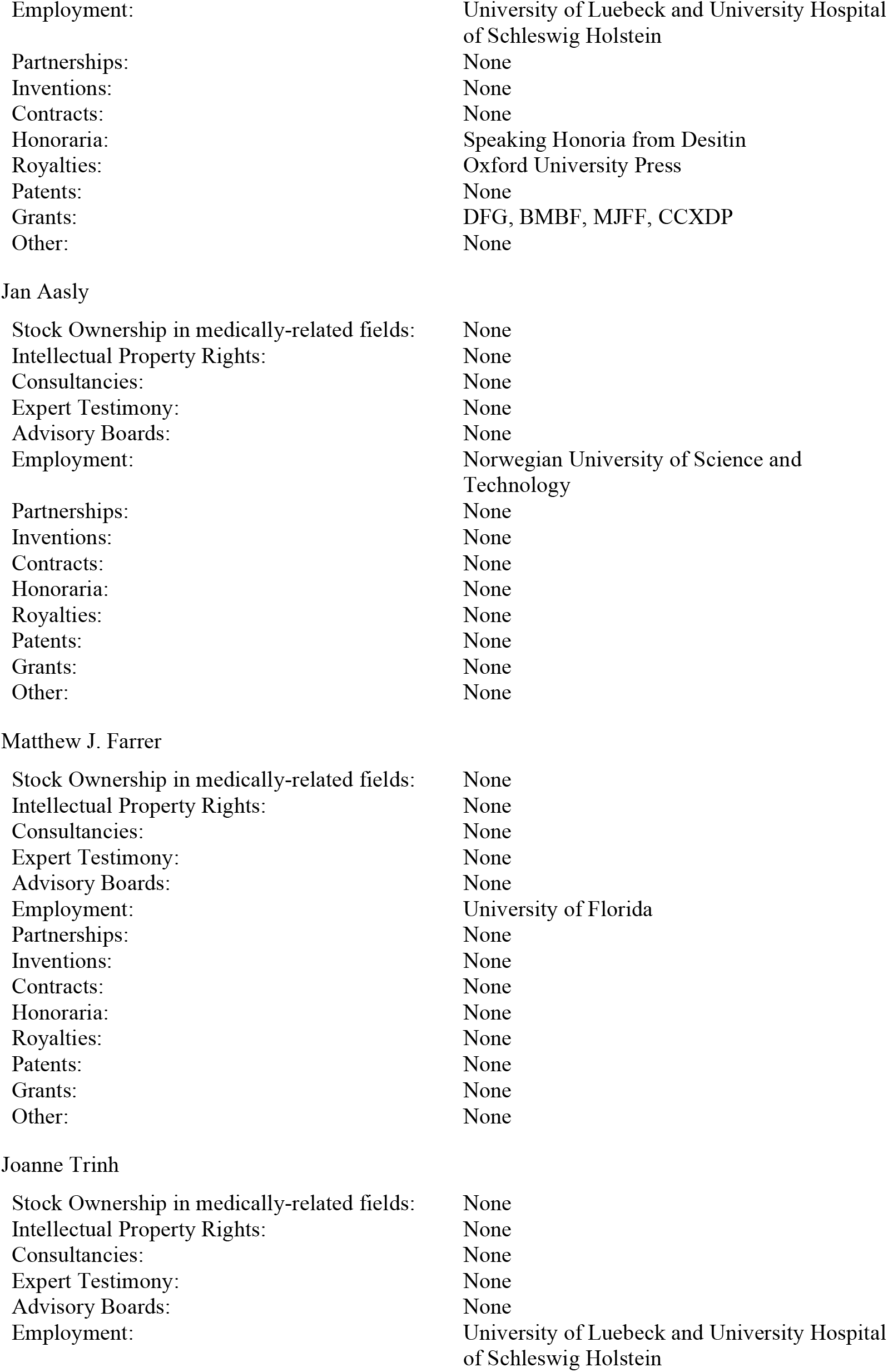
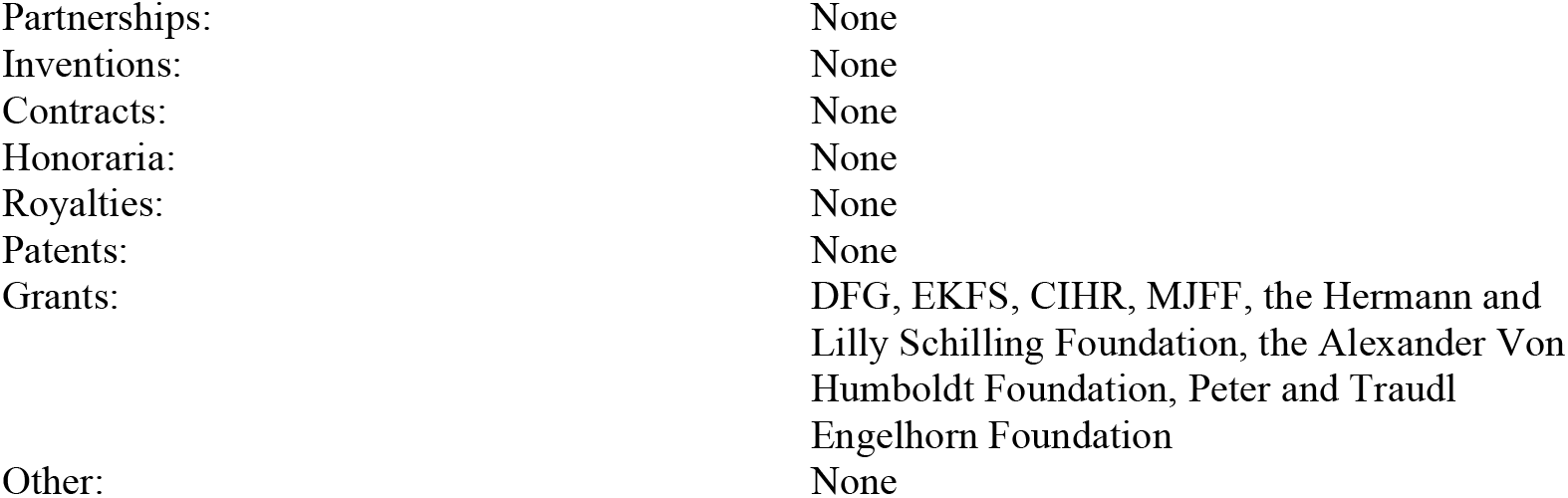

|  |  |
| --- | --- |
| Stock Ownership in medically-related fields: | None |
| Intellectual Property Rights: | None |
| Consultancies: | None |
| Expert Testimony: | None |
| Advisory Boards: | None |
| Employment: | University of Luebeck and University Hospital of Schleswig Holstein |
| Partnerships: | None |
| Inventions: | None |
| Contracts: | None |
| Honoraria: | None |
| Royalties: | None |
| Patents: | None |
| Grants: | DFG |
| Other: | None |
| Susen Schaake |  |
| Stock Ownership in medically-related fields: | None |
| Intellectual Property Rights: | None |
| Consultancies: | None |
| Expert Testimony: | None |
| Advisory Boards: | None |
| Employment: | University of Luebeck and University Hospital of Schleswig Holstein |
| Partnerships: | None |
| Inventions: | None |
| Contracts: | None |
| Honoraria: | None |
| Royalties: | None |
| Patents: | None |
| Grants: | None |
| Other: | None |
| Christoph Much |  |
| Stock Ownership in medically-related fields: | None |
| Intellectual Property Rights: | None |
| Consultancies: | None |
| Expert Testimony: | None |
| Advisory Boards: | None |
| Employment: | University of Luebeck and University Hospital of Schleswig Holstein |
| Partnerships: | None |
| Inventions: | None |
| Contracts: | None |
| Honoraria: | None |
| Royalties: | None |
| Patents: | None |
| Grants: | None |
| Other: | None |
| Christine Klein |  |
| Stock Ownership in medically-related fields: | None |
| Intellectual Property Rights: | None |
| Consultancies: | Centogene and Retromer Therapeutics |
| Expert Testimony: | None |
| Advisory Boards: | Else Kroener Fresenius Foundation |
| Employment: | University of Luebeck and University Hospital of Schleswig Holstein |
| Partnerships: | None |
| Inventions: | None |
| Contracts: | None |
| Honoraria: | Speaking Honoria from Desitin |
| Royalties: | Oxford University Press |
| Patents: | None |
| Grants: | DFG, BMBF, MJFF, CCXDP |
| Other: | None |
| Jan Aasly |  |
| Stock Ownership in medically-related fields: | None |
| Intellectual Property Rights: | None |
| Consultancies: | None |
| Expert Testimony: | None |
| Advisory Boards: | None |
| Employment: | Norwegian University of Science and Technology |
| Partnerships: | None |
| Inventions: | None |
| Contracts: | None |
| Honoraria: | None |
| Royalties: | None |
| Patents: | None |
| Grants: | None |
| Other: | None |
| Matthew J. Farrer |  |
| Stock Ownership in medically-related fields: | None |
| Intellectual Property Rights: | None |
| Consultancies: | None |
| Expert Testimony: | None |
| Advisory Boards: | None |
| Employment: | University of Florida |
| Partnerships: | None |
| Inventions: | None |
| Contracts: | None |
| Honoraria: | None |
| Royalties: | None |
| Patents: | None |
| Grants: | None |
| Other: | None |
| Joanne Trinh |  |
| Stock Ownership in medically-related fields: | None |
| Intellectual Property Rights: | None |
| Consultancies: | None |
| Expert Testimony: | None |
| Advisory Boards: | None |
| Employment: | University of Luebeck and University Hospital of Schleswig Holstein |
| Partnerships: | None |
| Inventions: | None |
| Contracts: | None |
| Honoraria: | None |
| Royalties: | None |
| Patents: | None |
| Grants: | DFG, EKFS, CIHR, MJFF, the Hermann and Lilly Schilling Foundation, the Alexander Von Humboldt Foundation, Peter and Traudl Engelhorn Foundation |
| Other: | None |

## Notes

### Competing Interest Statement

The authors have declared no competing interest.

### Author Declarations

Ethics Board of the University of Lübeck, Germany gave ethical approval for this work.

