## Supplementary tables and figures for "*GBA* in Parkinson’s disease: variant detection and pathogenicity scoring matters"

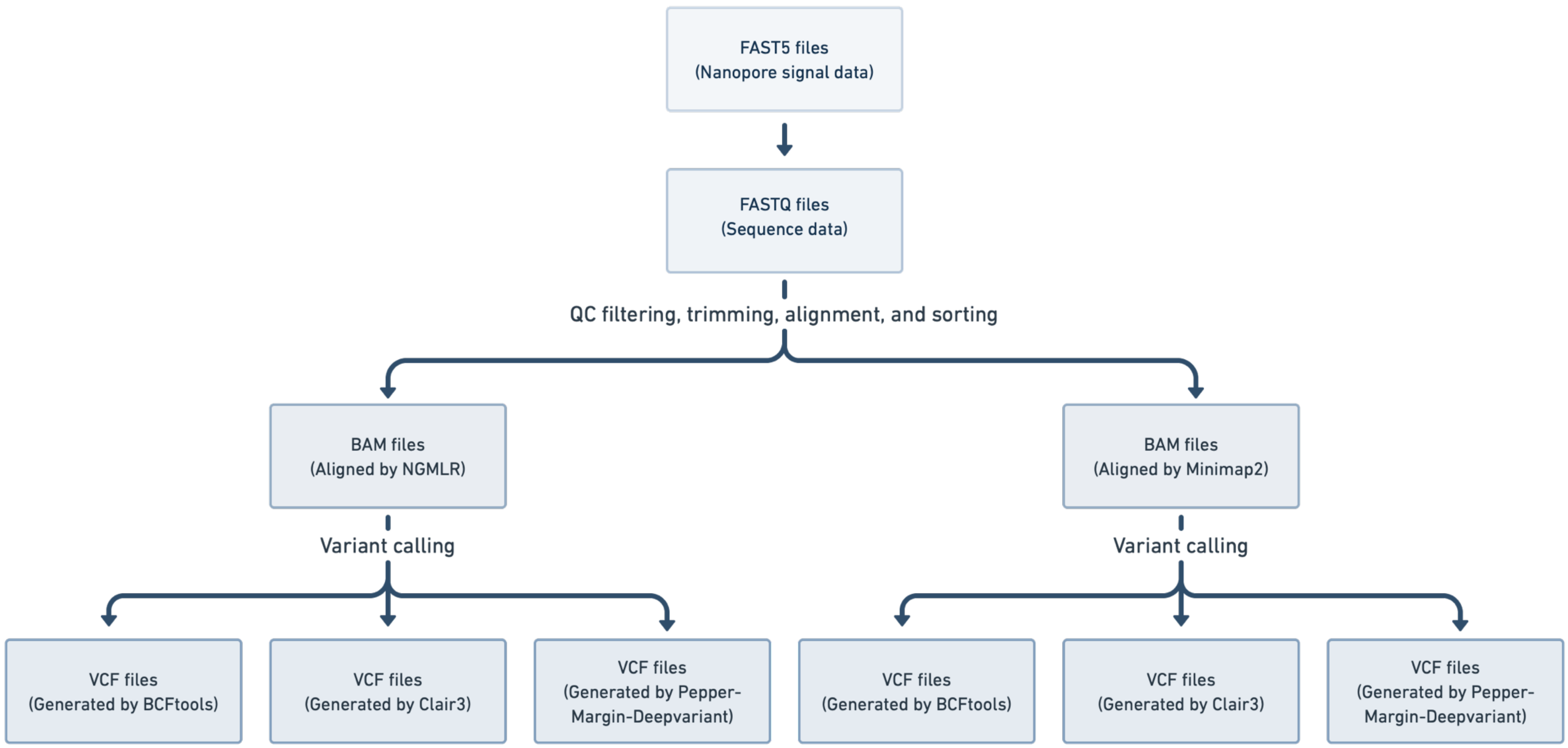


**Supplementary Figure 1.** Laboratory workflow data analysis pipeline of how the data was processed and which aligners (i.e. NGMLR and Minimap2) and variant callers (i.e. BCFtools, Clair3, Pepper-Margin-Deepvariant).

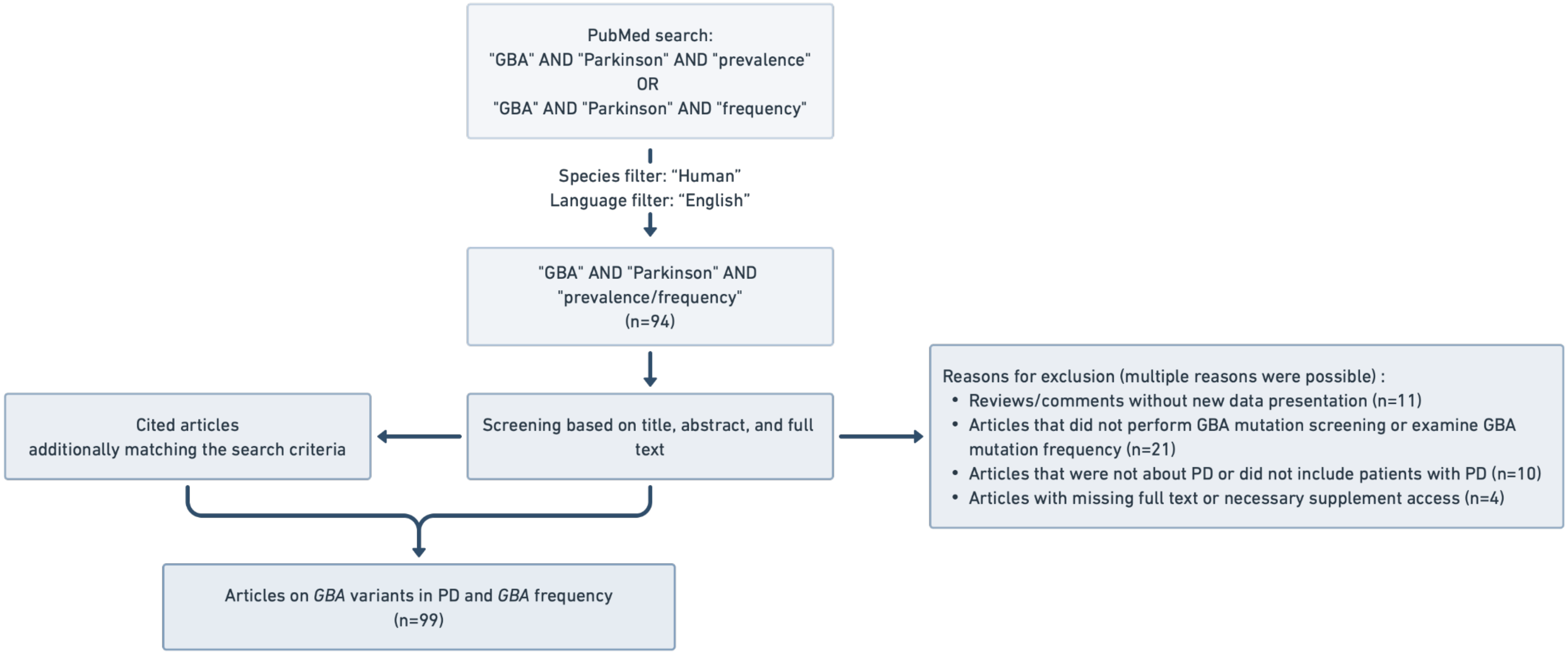


**Supplementary Figure 2.** Workflow of the literature search in PubMed. We searched for literature via PubMed that was published before August 4, 2022, using the search term "*GBA*" AND "Parkinson" AND "prevalence" OR "*GBA*" AND "Parkinson" AND "frequency", while setting the species filter to "Human" and the language filter to "English", resulting in 94 articles. These were screened based on the title, abstract and full text, excluding all articles not directly screening for variants in the *GBA* gene in patients with PD. Reasons for exclusion were reviews, or comments without new data (n=11), in this case, the original publications were examined, articles that did not perform *GBA* variant screening or examine *GBA* variant frequency in their study population (n=21), and articles that were not about PD or did not include patients with PD (n=10) (multiple reasons for exclusion were possible). Four more articles had to be excluded due to missing full articles or necessary supplement access. In addition, suited articles that were referenced in these articles were further included in the overview. In the end, 99 articles on *GBA* variant frequencies across populations were included in the overview.

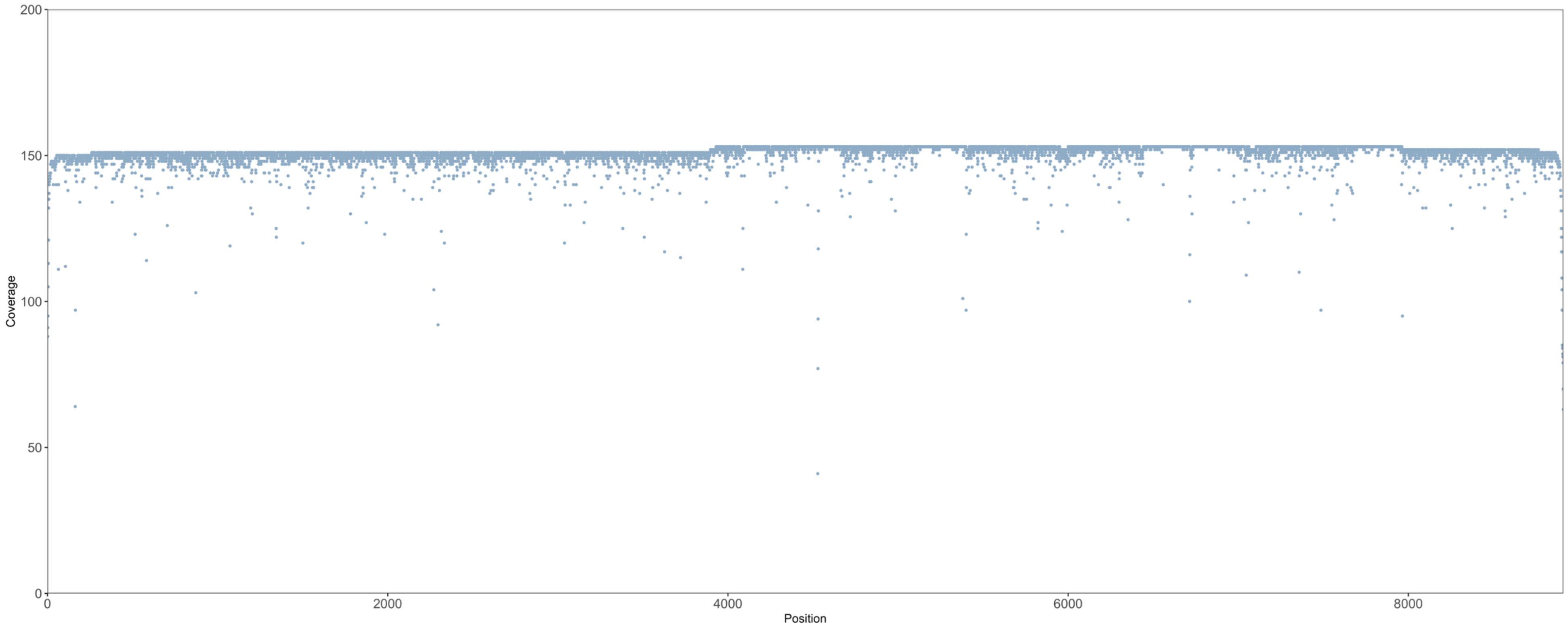


**Supplementary Figure 3.** Exemplary coverage plot. The coverage is shown over the full *GBA* amplicon (8.9 kb).

**Supplementary Table 1.** Overview over all rare *GBA* variants detected with Nanopore sequencing.

| Position chr1 (hg38) | REF | ALT | Region | SNP ID | Variant information *GBA* (NM_000157.4) | AA change | ACMG classification | ClinVar | SIFT | Poly-phen2 HDIV | Poly-phen2 HVAR | CADD raw | CADD phred | GERP++ | gnomAD (All) |
|---|---|---|---|---|---|---|---|---|---|---|---|---|---|---|---|
| 155232724 | C | T | intergenic | rs578095239 | . | . | Likely benign -1 points = 1 P - 2 B | . | . | . | . | . | . | . | 0.0002 |
| 155232838 | C | T | intergenic | rs561692357 | . | . | Uncertain significance 1 points = 1 P - 0 B | . | . | . | . | . | . | . | . |
| 155232871 | T | C | intergenic | rs529155642 | . | . | Likely benign -1 points = 1 P - 2 B | . | . | . | . | . | . | . | 0.0006 |
| 155232982 | G | A | intergenic | rs530421143 | . | . | Likely benign -1 points = 1 P - 2 B | . | . | . | . | . | . | . | 0.001 |
| 155233134 | C | T | intergenic | rs557608543 | . | . | Uncertain significance 0 points = 1 P - 1 B | . | . | . | . | . | . | . | 0.0005 |
| 155233268 | C | G | intergenic | rs367634752 | . | . | Likely benign -1 points = 1 P - 2 B | . | . | . | . | . | . | . | 0.0016 |
| 155233286 | C | T | intergenic | rs1042594015 | . | . | Likely benign -1 points = 1 P - 2 B | . | . | . | . | . | . | . | . |
| 155233287 | G | A | intergenic | rs368998334 | . | . | Likely benign -6 points = 0 P - 6 B | . | . | . | . | . | . | . | 0.0032 |
| 155233514 | A | G | downstream | rs2361530 | . | . | Likely benign -1 points = 1 P - 2 B | . | . | . | . | . | . | . | . |
| 155233517 | C | T | downstream | rs2361531 | . | . | Likely benign -1 points = 1 P - 2 B | . | . | . | . | . | . | . | . |
| 155233521 | T | A | downstream | rs2361532 | . | . | Likely benign -1 points = 1 P - 2 B | . | . | . | . | . | . | . | . |
| 155233531 | G | A | downstream | rs2361533 | . | . | Uncertain significance 0 points = 1 P - 1 B | . | . | . | . | . | . | . | . |
| 155233541 | G | T | downstream | rs4024047 | . | . | Likely benign -1 points = 1 P - 2 B | . | . | . | . | . | . | . | 0.00003238 |
| 155233543 | G | A | downstream | rs4024048 | . | . | Likely benign -1 points = 1 P - 2 B | . | . | . | . | . | . | . | 0.00003241 |
| 155233549 | C | T | downstream | rs4024049 | . | . | Likely benign -1 points = 1 P - 2 B | . | . | . | . | . | . | . | 0.00003258 |
| 155233612 | A | G | downstream | rs2142046 | . | . | Benign -10 points = 0 P - 10 B | . | . | . | . | . | . | . | 0.0024 |
| 155233639 | G | A | downstream | rs2142045 | . | . | Likely benign -1 points = 1 P - 2 B | . | . | . | . | . | . | . | 0.0005 |
| 155234414 | G | A | downstream | rs201209118 | . | . | Benign -10 points = 0 P - 10 B | . | . | . | . | . | . | . | 0 |
| 155234893 | A | G | UTR3 | rs368275143 | c.*102T>C | . | Likely benign -1 points = 1 P - 2 B | . | . | . | . | . | . | . | 0.0113 |
| 155234903 | C | T | UTR3 | rs708606 | c.*92G>A | . | Likely benign -1 points = 1 P - 2 B | . | . | . | . | . | . | . | 0.0022 |
| 155235104 | G | A | intronic | rs374690110 | c.1506-4C>T | . | Likely benign -1 points = 1 P - 2 B | . | . | . | . | . | . | . | 0.0002 |
| 155235206 | G | A | exonic | rs371779859 | c.1494C>T | p.Val498= | Likely benign -2 points = 1 P - 3 B | . | . | . | . | . | . | . | 0.00009689 |
| 155235222 | C | T | exonic | . | c.1478G>A | p.Gly493Asp | Uncertain significance 1 points = 1 P - 0 B | . | T | D | P | 4.834 | 24.8 | 3.16 | . |
| 155235252 | A | G | exonic | rs421016 | c.1448T>C | p.Leu483Pro | Pathogenic 11 points = 11 P – 0 B | Pathogenic; risk factor | D | P | P | 4.842 | 24.8 | 3.16 | 0.0007 |
| 155235302 | A | G | exonic | . | c.1398T>C | p.Ile466= | Likely benign -2 points = 1 P - 3 B | . | . | . | . | . | . | . | . |

| | | | | | | | | | | | | | | | |
|---|---|---|---|---|---|---|---|---|---|---|---|---|---|---|---|
| 155235344 | G | A | intronic | rs569191841 | c.1389-33C>T | . | Likely benign -1 points = 1 P - 2 B | . | . | . | . | . | . | . | 0.00003234 |
| 155235379 | A | G | intronic | rs2974924 | c.1389-68T>C | . | Likely benign -1 points = 1 P - 2 B | . | . | . | . | . | . | . | 0.0105 |
| 155235587 | C | T | intronic | rs12752133 | c.1388+94G>A | . | Likely benign -1 points = 1 P - 2 B | . | . | . | . | . | . | . | 0.0127 |
| 155235791 | G | A | exonic | rs201499639 | c.1278C>T | p.Pro426= | Likely benign -2 points = 1 P - 3 B | Likely benign | . | . | . | . | . | . | 0.0000323 |
| 155235843 | T | C | exonic | rs76763715 | c.1226A>G | p.Asn409Ser | Uncertain significance 5 points = 5 P – 0 B | Pathogenic/Likely pathogenic; risk factor | D | P | B | 3.202 | 22.7 | 3.53 | 0.0017 |
| 155235928 | G | C | intronic | rs41264925 | c.1225-84C>G | . | Likely benign -1 points = 1 P - 2 B | . | . | . | . | . | . | . | . |
| 155236050 | G | A | intronic | rs1036605613 | c.1224+195C>T | . | Uncertain significance 0 points = 1 P – 1 B | . | . | . | . | . | . | . | 0.00003232 |
| 155236246 | G | A | exonic | rs75548401 | c.1223C>T | p.Thr408Met | Uncertain significance 1 points = 1 P – 0 B | Uncertain significance (2); Benign (4); Likely benign (3) | T | B | B | 2.993 | 22.2 | 3.57 | 0.0076 |
| 155236294 | C | T | exonic | rs11558184 | c.1175G>A | p.Arg392Gln | Uncertain significance 1 points = 1 P – 0 B | . | T | P | B | 3.323 | 22.9 | 3.67 | 0.00003233 |
| 155236331 | C | T | exonic | rs781306264 | c.1138G>A | p.Ala380Thr | Uncertain significance 1 points = 1 P – 0 B | . | D | D | D | 6.843 | 33 | 3.67 | . |
| 155236366 | C | T | exonic | rs1064648 | c.1103G>A | p.Arg368His | Uncertain significance 1 points = 1 P – 0 B | . | T | B | B | 3.16 | 22.6 | 2.75 | . |
| 155236367 | G | A | exonic | rs374306700 | c.1102C>T | p.Arg368Cys | Uncertain significance 1 points = 1 P – 0 B | Likely pathogenic (2); Uncertain significance (2) | T | D | P | 6.006 | 27.9 | 3.67 | . |
| 155236376 | C | T | exonic | rs2230288 | c.1093G>A | p.Glu365Lys | Likely benign -1 points = 1 P – 2 B | Benign/Likely benign; risk factor | T | B | B | 2.173 | 17.33 | 3.67 | 0.0128 |
| 155236459 | T | C | exonic | rs1306645655 | c.1010A>G | p.Asp337Gly | Uncertain significance 1 points = 1 P – 0 B | . | D | D | D | 5.661 | 26.7 | 3.67 | . |
| 155236558 | C | T | intronic | rs772645370 | c.1000-89G>A | . | Likely benign -1 points = 1 P – 2 B | . | . | . | . | . | . | . | 0.0002 |
| 155236787 | G | A | intronic | rs531447697 | c.1000-318C>T | . | Likely benign -1 points = 1 P – 2 B | . | . | . | . | . | . | . | 0.00003263 |
| 155237162 | G | A | intronic | rs547873878 | c.999+179C>T | . | Likely benign -1 points = 1 P – 2 B | . | . | . | . | . | . | . | 0.0003 |
| 155237222 | A | C | intronic | rs946743963 | c.999+119T>G | . | Likely benign -1 points = 1 P – 2 B | . | . | . | . | . | . | . | 0.00003232 |
| 155237239 | G | A | intronic | rs72704130 | c.999+102C>T | . | Likely benign -1 points = 1 P – 2 B | . | . | . | . | . | . | . | 0.0138 |
| 155237265 | G | A | intronic | rs556277010 | c.999+76C>T | . | Likely benign -1 points = 1 P – 2 B | . | . | . | . | . | . | . | 0.00003232 |
| 155237412 | T | C | exonic | rs1057942 | c.928A>G | p.Ser310Gly | Uncertain significance 3 points = 3 P – 0 B | Pathogenic/Likely pathogenic | T | P | B | 1.77 | 14.81 | 3.51 | 0.00006461 |
| 155237438 | C | T | exonic | rs140955685 | c.902G>A | p.Arg301His | Uncertain significance 1 points = 1 P – 0 B | Uncertain significance | T | B | B | 2.204 | 17.54 | 2.59 | 0.0003 |
| 155237596 | A | T | intronic | rs140335079 | c.762-18T>G | . | Likely benign -1 points = 1 P – 2 B | . | . | . | . | . | . | . | 0.009 |
| 155237623 | C | G | intronic | rs377217353 | c.762-45G>C | . | Likely benign -1 points = 1 P – 2 B | . | . | . | . | . | . | . | 0.00003236 |
| 155237914 | A | G | intronic | rs549565365 | c.761+220T>C | . | Likely benign -1 points = 1 P – 2 B | . | . | . | . | . | . | . | 0.0045 |
| 155238057 | G | T | intronic | rs183540501 | c.761+77C>A | . | Likely benign -1 points = 1 P – 2 B | . | . | . | . | . | . | . | 0.0031 |
| 155238175 | G | A | exonic | rs376613535 | c.720C>T | p.Pro240= | Likely benign -2 points = 1 P – 3 B | . | . | . | . | . | . | . | 0.00006465 |
| 155238265 | G | A | exonic | rs201615998 | c.630C>T | p.Pro210= | Likely benign | . | . | . | . | . | . | . | . |

| | | | | | | | | | | | | | | | |
|---|---|---|---|---|---|---|---|---|---|---|---|---|---|---|---|
| | | | | | | | -2 points = 1 P – 3 B | | | | | | | | |
| 155238570 | C | G | exonic | rs147138516 | c.535G>C | p.Asp179His | Uncertain significance 1 points = 1 P – 0 B | Likely pathogenic (1); Uncertain significance (2) | T | P | P | 0.611 | 8.229 | 2.62 | 0.00009723 |
| 155238629 | C | T | exonic | rs79653797 | c.476G>A | p.Arg159Gln | Uncertain significance 3 points = 3 P – 0 B | Pathogenic/Likely pathogenic | D | D | D | 6.336 | 29.3 | 3.55 | . |
| 155238630 | G | A | exonic | rs439898 | c.475C>T | p.Arg159Trp | Uncertain significance 3 points = 3 P – 0 B | Pathogenic | D | D | D | 6.19 | 28.6 | 3.55 | 0.00003238 |
| 155238631 | G | A | exonic | rs147411159 | c.474C>T | p.Ile158= | Likely benign -2 points = 1 P – 3 B | Uncertain significance (1); Likely benign (3) | . | . | . | . | . | . | 0.0006 |
| 155238833 | A | G | intronic | rs188328778 | c.455-183T>C | . | Likely benign -1 points = 1 P – 2 B | . | . | . | . | . | . | . | 0.0119 |
| 155238857 | G | A | intronic | rs1042674060 | c.455-207C>T | . | Likely benign -1 points = 1 P – 2 B | . | . | . | . | . | . | . | 0.0001 |
| 155238927 | A | G | intronic | rs778649863 | c.455-277T>C | . | Likely benign -1 points = 1 P – 2 B | . | . | . | . | . | . | . | . |
| 155238984 | C | G | intronic | rs752258174 | c.455-334G>C | . | Likely benign -1 points = 1 P – 2 B | . | . | . | . | . | . | . | . |
| 155238985 | G | A | intronic | rs951266434 | c.455-335C>T | . | Likely benign -1 points = 1 P – 2 B | . | . | . | . | . | . | . | . |
| 155239079 | G | A | intronic | rs572108051 | c.455-429C>T | . | Likely benign -1 points = 1 P – 2 B | . | . | . | . | . | . | . | 0 |
| 155239287 | C | T | intronic | rs1005434278 | c.454+329G>A | . | Likely benign -1 points = 1 P – 2 B | . | . | . | . | . | . | . | 0.00003262 |
| 155239509 | C | T | intronic | rs570088632 | c.454+107G>A | . | Likely benign -1 points = 1 P – 2 B | . | . | . | . | . | . | . | . |
| 155239633 | G | T | exonic | rs758447515 | c.437C>A | p.Ser146Ter | Likely pathogenic 9 points = 9 P – 0 B | . | . | . | . | 10.665 | 36 | 3.25 | . |
| 155239858 | C | A | intronic | rs369792423 | c.307+28G>T | . | Likely benign -1 points = 1 P – 2 B | . | . | . | . | . | . | . | 0.00006462 |
| 155239939 | C | T | exonic | rs77829017 | c.254G>A | p.Gly85Glu | Uncertain significance 1 points = 1 P – 0 B | Pathogenic | D | D | D | 4.841 | 24.8 | 3.46 | . |
| 155239961 | G | A | exonic | rs146774384 | c.232C>T | p.Arg78Cys | Uncertain significance 1 points = 1 P – 0 B | . | T | D | P | 4.822 | 24.8 | 3.46 | 0.00009697 |
| 155240072 | G | C | exonic | . | c.121C>G | p.Arg41Gly | Uncertain significance 1 points = 1 P – 0 B | . | T | B | B | 0.931 | 10.26 | 3.41 | . |
| 155240122 | T | G | intronic | rs199565854 | c.116-45A>C | . | Likely benign -1 points = 1 P – 2 B | . | . | . | . | . | . | . | 0.0003 |
| 155240171 | C | T | intronic | rs114217696 | c.116-94G>A | . | Likely benign -1 points = 1 P – 2 B | . | . | . | . | . | . | . | 0.008 |
| 155240336 | C | T | intronic | rs142348200 | c.116-259G>A | . | Likely benign -1 points = 1 P – 2 B | . | . | . | . | . | . | . | 0.0012 |
| 155240779 | T | C | intronic | rs2361534 | c.28-62A>G | . | Likely benign -1 points = 1 P – 2 B | . | . | . | . | . | . | . | 0.001 |
| 155240816 | G | A | intronic | rs940168433 | c.28-99C>T | . | Likely benign -1 points = 1 P – 2 B | . | . | . | . | . | . | . | . |
| 155241114 | C | T | UTR5 | rs1141801 | c.-2G>T | . | Likely benign -1 points = 1 P – 2 B | . | . | . | . | . | . | . | . |
| 155241127 | T | C | UTR5 | rs41264927 | c.-15A>G | . | Likely benign -1 points = 1 P – 2 B | . | . | . | . | . | . | . | 0.0012 |
| 155241257 | C | T | intronic | rs371157845 | NM_001005742.3):c.-49-96G>A | . | Likely benign -1 points = 1 P – 2 B | . | . | . | . | . | . | . | . |
| 155241315 | T | C | intronic | rs188978150 | NM_001005742.3):c.-49-154A>G | . | Likely benign -1 points = 1 P – 2 B | Uncertain Significance | . | . | . | . | . | . | 0.0086 |

Variants with a gnomAD frequency >2% and without information on SNP ID or amino acid change were excluded. Pathogenicity scores were used from ACMG, Varsome, Clinvar, SIFT, Polphen2, CADD, and GERP++. Variants that were Sanger sequenced are highlighted in gray.

**Supplementary Table 2.** Rare *GBA* variants (predicted as "pathogenic"/"likely pathogenic"/"uncertain significance") sequenced with the Oxford Nanopore and confirmed with Sanger sequencing.

| ***GBA* variant** | **cDNA transcript *GBA*(NM_000157.4)** | **Exon number** | **PD Cases (n=462)** | **Controls (n=367)** |
|---|---|---|---|---|
| p.R78C | c.232C>T | 3 | 2 | 0 |
| p.S146X | c.437C>A | 4 | 1 | 0 |
| p.R159W | c.475C>T | 5 | 12 | 1 |
| p.R301H | c.902G>A | 7 | 0 | 1 |
| p.S310G | c.928A>G | 7 | 1 | 1 |
| p.D337G | c.1010A>G | 8 | 2 | 1 |
| p.E365K | c.1093G>A | 8 | 33 | 11 |
| p.R368C | c.1102C>T | 8 | 2 | 0 |
| p.A380T | c.1138G>A | 8 | 1 | 0 |
| p.T408M | c.1223C>T | 8 | 10 | 13 |
| p.N409S | c.1226A>G | 9 | 13 | 4 |
| p.L483P | c.1448T>C | 10 | 6 | 0 |
| p.G493D | c.1478G>A | 10 | 1 | 0 |

**Supplementary Table 3.** Publications on *GBA* variants and frequencies in patients with PD included in PubMed.

| Authors, Year (PMID) | n (PD/control) | Population/ Region | Method | *GBA* variants screened* | *GBA* variants found* | Counts/Frequency* |
|---|---|---|---|---|---|---|
| Toft et al., 2006[1] (16476943) | 311/474 | Norwegian | Variant screening | L444P, N370S | L444P, N370S | L444P: PD: 3/311 (0.96%), Controls: 1/474 (0.21%)<br>N370S: PD: 4/311 (1.29%), Controls: 7/474 (1.48%) |
| Lunde et al., 2018[2] (29792872) | 442/419 | Norwegian | Genotyping | N370S, T369M, E326K, V460L, Y135C, L444P | N370S, T369M, E326K, V460L, Y135C, L444P | All: PD: 53/442 (12.0%), Controls: 29/419 (6.9%)<br>N370S: PD: 1/442 (0.2%), Controls: 1/419 (0.2%)<br>T369M: PD: 7/442 (1.7%), Controls: 16/419 (3.6%)<br>E326K: PD: 18/442 (4.3%), Controls: 29/419 (6.6%)<br>V460L: PD: 1/442 (0.2%), Controls: 1/419 (0.2%)<br>Y135C: PD: 0/442 (0%), Controls: 1/419 (0.2%)<br>L444P: PD: 2/442 (0.5%), Controls: 6/419 (1.4%) |
| Berge-Seidl et al., 2017[3] (28830825) | 1152/713 | Scandinavian | Targeted deep sequencing; genotyping | All *GBA* exons | E326K, T369M, N370S, R463C, IVS3+1G>A, V457A, G377D, W357R | E326K: PD: 20/366 (5.46%)<br>T369M: PD: 13/366 (3.55%)<br>N370S: PD: 1/366 (0.27%)<br>R463C: PD: 1/366 (0.27%)<br>IVS3+1G>A: PD: 1/366 (0.27%)<br>V457A: PD: 1/366 (0.27%)<br>G377D: PD: 1/366 (0.27%)<br>W357R: PD: 1/366 (0.27%) |
| Ran et al., 2022[4] (35779693) | 1131/1594 | Swedish | Pyrosequencing for genotyping of T369M | T369M | T369M | T369M: PD: 47/1091 (4.31%), Controls: 50/1474 (3.39%) |
| Ran et al., 2016[5] (27255555) | 1625/2025 | Swedish | Genotyping by pyrosequencing of E326K, N370S, and L444P | E326K, N370S, L444P | E326K, N370S, L444P | E326K: PD: 90/1625 (5.54%), Controls: 65/2025 (3.21%)<br>N370S: PD: 10/1625 (0.62%), Controls: 2/2025 (0.10%)<br>L444P: PD: 35/1625 (2.15%), Controls: 65/2025 (0.15%) |
| Ylönen et al., 2017[6] (29029963) | 862/403 | Finnish | Variant screening for N370S and L444P; Whole exome sequencing in 225 EOPD cases | All *GBA* exons; N370S, L444P | N370S, L444P | N370S: PD: 4/862 (0.5%), Controls: 1/403 (0.2%)<br>L444P: PD: 17/862 (2.0%), Controls: 2/403 (0.5%) |
| Muldmaa et al., 2021[7] (32740907) | 189/158 | Estonian | Next-generation sequencing | NA | L444P, T369M, E326K, L276I, E10X | All: PD: 19/189 (10.1%), Controls: 6/158 (3.8%)<br>*GBA*-related risk variants: 18/189 (9.5%)<br>L444P: PD: 1/189 (0.5%), Controls: 0/158 (0%)<br>T369M: PD: 10/189 (5.3%), Controls: 3/158 (1.9%)<br>E326K: PD: 6/189 (3.2%), Controls: 3/158 (1.9%)<br>L276I: PD: 1/189 (0.5%), Controls: 0/158 (0%)<br>E10X: PD: 1/189 (0.5%), Controls: 0/158 (0%) |
| Neumann et al., 2009[8] (19286695) | 790/257 | British | DNA sequencing of full *GBA* gene | All *GBA* exons and the flanking introns | L483P, D482N, R502C, RecNciI (L483P, A495P, V499V), RecA456P (L483P, A495P), N409S, D448H, D419A, N421PfsX4, R296Q, G232E, R170C, K46E, V497L | All: PD: 33/790 (4.18%), Controls: 3/257 (1.17%)<br>L483P: PD: 11/790 (1.39%), Controls: 0/257 (0%)<br>D482N: PD: 1/790 (0.13%), Controls: 0/257 (0%)<br>R502C: PD: 3/790 (0.38%), Controls: 0/257 (0%)<br>RecNciI (L483P, A495P, V499V): PD: 2/790 (0.25%), Controls: 0/257 (0%)<br>RecA456P (L483P, A495P): PD: 1/790 (0.13%), Controls: 0/257 (0%)<br>N409S: PD: 8/790 (1.01%), Controls: 1/257 (0.39%)<br>D448H: PD: 1/790 (0.13%), Controls: 0/257 (0%)<br>D419A: PD: 1/790 (0.13%), Controls: 0/257 (0%)<br>N421PfsX4: PD: 1/790 (0.13%), Controls: 0/257 (0%)<br>R296Q: PD: 1/790 (0.13%), Controls: 1/257 (0.39%)<br>G232E: PD: 1/790 (0.13%), Controls: 0/257 (0%)<br>R170C: PD: 1/790 (0.13%), Controls: 0/257 (0%) |

| | | | | | | |
|---|---|---|---|---|---|---|
| | | | | | | K46E: PD: 1/790 (0.13%), Controls: 0/257 (0%)<br>V497L: PD: 0/790 (0%), Controls: 1/257 (0.39%) |
| Winder-Rhodes et al., 2013[9]<br>(23413260) | 259/0 | British (2 South Asian) | DNA sequencing of all *GBA* exons | All *GBA* exons | L444P, N370S, N462K, R463C, R257Q, E326K, T369M, E388K, L119L | All: PD: 9/259 (3.5%)<br>L444P: PD: 3/259 (1.2%)<br>N370S: PD: 3/259 (1.2%)<br>N462K: PD: 1/259 (0.4%)<br>R463C: PD: 1/259 (0.4%)<br>R257Q: PD: 1/259 (0.4%)<br>E326K: PD: 8/259 (3.1%)<br>T369M: PD: 5/259 (1.9%)<br>E388K: PD: 1/259 (0.4%)<br>L119L: PD: 1/259 (0.4%) |
| Duran et al., 2013[10]<br>(23225227) | 185/283 | UK Caucasian | Sanger sequencing of the full *GBA* gene | Full *GBA* gene | N370S, L444P, RecNciI (L444P+A456P+V460V), R463C, E326K, IVS2+1, R131C, W184R, N188S, H255Q, R257Q, D409H, RecTL, E388K, G113A, T369M, S465P, L(−14)V, V172L, S177T, L217P, L317L, L354P, V375G, IVS10-4 C>T, IVS10-12 C>T, E340A, V458L | All: PD: 48/185 (25.94%), Controls: 12/283 (4.24%)<br>All pathogenic: PD: 37/185 (20%), Controls: 9/283 (3.18%)<br>N370S: PD: 5/185 (2.70%), Controls: 1/283 (0.35%)<br>L444P: PD: 2/185 (1.08%), Controls: 0/283 (0%)<br>RecNciI (L444P+A456P+V460V): PD: 3/185 (1.62%), Controls: 0/283 (0%)<br>R463C: PD: 3/185 (1.62%), Controls: 0/283 (0%)<br>E326K: PD: 14/185 (7.57%), Controls: 7/283 (2.47%), 13/485 (2.68%)<br>IVS2+1: PD: 1/185 (0.54%), Controls: 0/283 (0%)<br>R131C: PD: 2/185 (1.08%), Controls: 0/283 (0%)<br>W184R: PD: 1/185 (0.54%), Controls: 0/283 (0%)<br>N188S: PD: 1/185 (0.54%), Controls: 0/283 (0%)<br>H255Q: PD: 1/185 (0.54%), Controls: 0/283 (0%)<br>R257Q: PD: 1/185 (0.54%), Controls: 1/283 (0.35%)<br>D409H: PD: 2/185 (1.08%), Controls: 0/283 (0%)<br>RecTL: PD: 1/185 (0.54%), Controls: 0/283 (0%)<br>E388K: PD: 1/185 (0.54%), Controls: 0/283 (0%)<br>G113A: PD: 2/185 (1.08%), Controls: 0/283 (0%)<br>T369M: PD: 1/185 (0.54%), Controls: 1/283 (0.35%)<br>S465P: PD: 1/185 (0.54%), Controls: 0/283 (0%)<br>L(−14)V: PD: 1/185 (0.54%), Controls: 0/283 (0%)<br>V172L: PD: 2/185 (1.08%), Controls: 0/283 (0%)<br>S177T: PD: 1/185 (0.54%), Controls: 0/283 (0%)<br>L217P: PD: 1/185 (0.54%), Controls: 0/283 (0%)<br>L317L: PD: 1/185 (0.54%), Controls: 0/283 (0%)<br>L354P: PD: 1/185 (0.54%), Controls: 0/283 (0%)<br>V375G: PD: 1/185 (0.54%), Controls: 0/283 (0%)<br>IVS10-4 C>T: PD: 1/185 (0.54%), Controls: 0/283 (0%)<br>IVS10-12 C>T: PD: 1/185 (0.54%), Controls: 0/283 (0%)<br>E340A: PD: 0/185 (0%), Controls: 1/283 (0.35%)<br>V458L: PD: 0/185 (0%), Controls: 1/283 (0.35%) |
| Olszewska et al., 2020[11]<br>(32714263) | 314/96 (friends or spouses) | Irish | DNA sequencing of all *GBA* exons | All *GBA* exons | T408M, E365K, F255Y, N409S, D448H, L483P, A495P, V499V, G416C, G234E, R301H, R368C | T408M: PD: 6/314 (1.91%), Controls: 4/96 (4.17%)<br>E365K: PD: 13/314 (4.14%), Controls: 4/96 (4.17%)<br>F255Y: PD: 1/314 (0.32%), Controls: 0/96 (0%)<br>N409S: PD: 3/314 (0.96%), Controls: 0/96 (0%)<br>D448H: PD: 1/314 (0.32%), Controls: 0/96 (0%)<br>L483P: PD: 3/314 (0.96%), Controls: 0/96 (0%)<br>A495P: PD: 3/314 (0.96%), Controls: 0/96 (0%)<br>V499V: PD: 3/314 (0.96%), Controls: 0/96 (0%)<br>G416C: PD: 1/314 (0.32%), Controls: 0/96 (0%)<br>G234E: PD: 1/314 (0.32%), Controls: 0/96 (0%)<br>R301H: PD: 1/314 (0.32%), Controls: 0/96 (0%)<br>R368C: PD: 1/314 (0.32%), Controls: 0/96 (0%) |

| | | | | | | |
|---|---|---|---|---|---|---|
| Crosiers et al., 2016[12] (27397011) | 266/536 | Flanders-Belgian | In-depth Sanger sequencing of all *GBA* exons | All *GBA* exons | D179H, Q256SfsX9, L363P, N409S, L483P, RecNcil (L483P-A495S-V499V), E365K, T408M, G39R, H529R | All rare: PD: 12/266 (4.5%), Controls: 2/536 (0.37%)<br>D179H: PD: 1/266 (0.4%), Controls: 0/536 (0%)<br>Q256SfsX9: PD: 1/266 (0.4%), Controls: 0/536 (0%)<br>L363P: PD: 1/266 (0.4%), Controls: 0/536 (0%)<br>N409S: PD: 3/266 (1.1%), Controls: 1/536 (0.2%)<br>L483P: PD: 3/266 (1.1%), Controls: 1/536 (0.2%)<br>RecNcil (L483P-A495S-V499V): PD: 1/266 (0.4%), Controls: 0/536 (0%)<br>E365K: PD: 12/266 (4.5%), Controls: 15/536 (2.8%)<br>T408M: PD: 3/266 (1.1%), Controls: 11/536 (2.0%)<br>G39R: PD: 1/266 (0.4%), Controls: 0/536 (0%)<br>H529R: PD: 1/266 (0.4%), Controls: 0/536 (0%) |
| den Heijer et al., 2020[13] (32618053) | 3402/655 | Dutch | Next-generation sequencing of full *GBA* gene | Full *GBA* gene | E-30Gfs*8, L-24S, L-24S+S23G, Q-7R, C18*, R39C, S45Rfs*15, R120W, D140H, R170H, A190T, G202R, F216Y, G250S, H255Q, I260T, L324P, G325R, E326K, R329C, W348G, Q350H, T369M, N370S, V375G, D380Y, E388K, N392S, D409H, L444P, D453L, V460M, R463P, S484L, S488T, H490R, L268=, S271G, A456P, V460=, S-1T, V459=, R496H, G390E, V17=, T61=, I119=, I130=, Q143=, G193=, G195=, G344=, T369=, P452=, c.762-5G>A, c.1000-4G>T | E-30Gfs*8: PD: 1/3402 (0.03%), Controls: 0/655 (0%)<br>L-24S: PD: 1/3402 (0.03%), Controls: 0/655 (0%)<br>L-24S+S23G: PD: 1/3402 (0.03%), Controls: 0/655 (0%)<br>Q-7R: PD: 2/3402 (0.06%), Controls: 0/655 (0%)<br>C18*: PD: 1/3402 (0.03%), Controls: 0/655 (0%)<br>R39C: PD: 1/3402 (0.03%), Controls: 0/655 (0%)<br>S45Rfs*15: PD: 1/3402 (0.03%), Controls: 0/655 (0%)<br>R120W: PD: 5/3402 (0.15%), Controls: 0/655 (0%)<br>D140H: PD: 84/3402 (2.47%), Controls: 6/655 (0.92%)<br>R170H: PD: 2/3402 (0.06%), Controls: 0/655 (0%)<br>A190T: PD: 1/3402 (0.03%), Controls: 0/655 (0%)<br>G202R: PD: 1/3402 (0.03%), Controls: 0/655 (0%)<br>F216Y: PD: 1/3402 (0.03%), Controls: 0/655 (0%)<br>G250S: PD: 1/3402 (0.03%), Controls: 0/655 (0%)<br>H255Q: PD: 2/3402 (0.06%), Controls: 0/655 (0%)<br>I260T: PD: 2/3402 (0.06%), Controls: 0/655 (0%)<br>L324P: PD: 2/3402 (0.06%), Controls: 1/655 (0.15%)<br>G325R: PD: 1/3402 (0.03%), Controls: 0/655 (0%)<br>E326K: PD: 314/3402 (9.23%), Controls: 18/655 (2.75%)<br>R329C: PD: 2/3402 (0.06%), Controls: 0/655 (0%)<br>W348G: PD: 1/3402 (0.03%), Controls: 0/655 (0%)<br>Q350H: PD: 1/3402 (0.03%), Controls: 1/655 (0.15%)<br>T369M: PD: 98/3402 (2.88%), Controls: 12/655 (1.83%)<br>N370S: PD: 32/3402 (0.94%), Controls: 2/655 (0.31%)<br>V375G: PD: 1/3402 (0.03%), Controls: 0/655 (0%)<br>D380Y: PD: 1/3402 (0.03%), Controls: 0/655 (0%)<br>E388K: PD: 3/3402 (0.09%), Controls: 0/655 (0%)<br>N392S: PD: 1/3402 (0.03%), Controls: 0/655 (0%)<br>D409H: PD: 1/3402 (0.03%), Controls: 0/655 (0%)<br>L444P: PD: 26/3402 (0.76%), Controls: 0/655 (0%)<br>D453L: PD: 5/3402 (0.15%), Controls: 0/655 (0%)<br>V460M: PD: 1/3402 (0.03%), Controls: 0/655 (0%)<br>R463P: PD: 2/3402 (0.06%), Controls: 1/655 (0.15%)<br>S484L: PD: 1/3402 (0.03%), Controls: 0/655 (0%)<br>S488T: PD: 1/3402 (0.03%), Controls: 0/655 (0%)<br>H490R: PD: 1/3402 (0.03%), Controls: 0/655 (0%)<br>L268=+S271G+D409H: PD: 1/3402 (0.03%), Controls: 0/655 (0%)<br>RecTL (D409H+L444P+A456P+V460=): PD: 1/3402 (0.03%), Controls: 0/655 (0%)<br>RecNciI (L444P+A456P+V460=): PD: 4/3402 (0.12%), Controls: 0/655 (0%)<br>S-1T: PD: 1/3402 (0.03%), Controls: 0/655 (0%)<br>V459=: PD: 5/3402 (0.15%), Controls: 0/655 (0%)<br>R496H: PD: 1/3402 (0.03%), Controls: 0/655 (0%)<br>G390E: PD: 1/3402 (0.03%), Controls: 1/655 (0.15%)<br>V17=: PD: 0/3402 (0%), Controls: 1/655 (0.15%)<br>T61=: PD: 1/3402 (0.03%), Controls: 0/655 (0%)<br>I119=: PD: 5/3402 (0.15%), Controls: 0/655 (0%) |

| | | | | | | |
|---|---|---|---|---|---|---|
| | | | | | | I130=: PD: 1/3402 (0.03%), Controls: 0/655 (0%)<br>Q143=: PD: 1/3402 (0.03%), Controls: 0/655 (0%)<br>G193=: PD: 1/3402 (0.03%), Controls: 1/655 (0.15%)<br>G195=: PD: 1/3402 (0.03%), Controls: 0/655 (0%)<br>G344=: PD: 1/3402 (0.03%), Controls: 0/655 (0%)<br>T369=: PD: 2/3402 (0.06%), Controls: 0/655 (0%)<br>P452=: PD: 1/3402 (0.03%), Controls: 0/655 (0%)<br>V460=: PD: 6/3402 (0.18%), Controls: 0/655 (0%)<br>c.762-5G>A: PD: 1/3402 (0.03%), Controls: 0/655 (0%)<br>c.1000-4G>T: PD: 0/3402 (0%), Controls: 1/655 (0.15%) |
| Anheim et al., 2012[14] (22282650) | 525/71 (relatives) | French (88%) | DNA sequencing of all *GBA* exons | All *GBA* exons | N370S, L444P, F246L, G202R, R120W, R463C, R463H, S125N, S173SfsX50, S364N, T323I, Y304C, 1263-1217del55bp, E326K | All: PD (probands): 24/525 (4.6%)<br>N370S: PD (probands): 10/525 (1.9%), Relatives with PD: 9/32 (28.1%), Relatives without PD: 12/71 (16.9%)<br>L444P: PD (probands): 5/525 (1.0%), Relatives with PD: 6/32 (18.8%), Relatives without PD: 5/71 (7.0%)<br>F246L: PD (probands): 1/525 (0.2%), Relatives with PD: 0/32 (0%), Relatives without PD: 0/71 (0%)<br>G202R: PD (probands): 1/525 (0.2%), Relatives with PD: 0/32 (0%) Relatives without PD: 0/71 (0%)<br>R120W: PD (probands): 1/525 (0.2%), Relatives with PD: 1/32 (3.1%), Relatives without PD: 0/71 (0%)<br>R463C: PD (probands): 1/525 (0.2%), Relatives with PD: 1/32 (3.1%), Relatives without PD: 1/71 (1.4%)<br>R463H: PD (probands): 1/525 (0.2%), Relatives with PD: 3/32 (9.4%), Relatives without PD: 3/71 (4.2%)<br>S125N: PD (probands): 1/525 (0.2%), Relatives with PD: 1/32 (3.1%), Relatives without PD: 0/71 (0%)<br>S173SfsX50: PD (probands): 1/525 (0.2%), Relatives with PD: 1/32 (3.1%), Relatives without PD: 3/71 (4.2%)<br>S364N: PD (probands): 1/525 (0.2%), Relatives with PD: 1/32 (3.1%), Relatives without PD: 0/71 (0%)<br>T323I: PD (probands): 1/525 (0.2%), Relatives with PD:1/32 (3.1%), Relatives without PD: 1/71 (1.4%)<br>Y304C: PD (probands): 1/525 (0.2%), Relatives with PD: 2/32 (6.3%), Relatives without PD: 6/71 (8.5%)<br>1263-1217del55bp: PD (probands): 1/525 (0.2%), Relatives with PD: 1/32 (3.1%), Relatives without PD: 0/71 (0%)<br>E326K: PD (probands): 0/525 (0%), Relatives with PD: 1/32 (3.1%), Relatives without PD: 0/71 (0%) |
| Lesage et al., 2011[15] (20947659) | 1130/391 (mainly spouses) | French (89%) | DNA sequencing of full *GBA* gene | *GBA* exons and flanking introns | K(227)R, K79M, G80R, I119L, R120W, S125N, R131C, S173SfsX50, G202R, P246L, Y304C, T323I, R329C, S364N, N370S, G377S, E388K, D409H, L444P, P452L, R463C, R463H, G113A, A446A, RecΔ5, RecNciI (L444P+A456P+V460V), RecA456P (L444P+A456P), c.1263del+RecTL (c.1263–1317del+D409H+L444P+A456P+V460V), D140H, E326K, A190A, Y313Y | K(227)R: PD: 1/1130 (0.09%), Controls: 0/391 (0%)<br>K79M: PD: 0/1130 (0%), Controls: 1/391 (0.26%)<br>G80R: PD: 1/1130 (0.09%), Controls: 0/391 (0%)<br>I119L: PD: 1/1130 (0.09%), Controls: 0/391 (0%)<br>R120W: PD: 1/1130 (0.09%), Controls: 0/391 (0%)<br>S125N: PD: 1/1130 (0.09%), Controls: 0/391 (0%)<br>R131C: PD: 1/1130 (0.09%), Controls: 0/391 (0%)<br>S173SfsX50: PD: 1/1130 (0.09%), Controls: 0/391 (0%)<br>G202R: PD: 2/1130 (0.18%), Controls: 0/391 (0%)<br>P246L: PD: 1/1130 (0.09%), Controls: 0/391 (0%)<br>Y304C: PD: 1/1130 (0.09%), Controls: 0/391 (0%)<br>T323I: PD: 1/1130 (0.09%), Controls: 0/391 (0%)<br>R329C: PD: 2/1130 (0.18%), Controls: 0/391 (0%)<br>S364N: PD: 1/1130 (0.09%), Controls: 0/391 (0%)<br>N370S: PD: 37/1130 (3.27%), Controls: 2/391 (0.51%)<br>G377S: PD: 1/1130 (0.09%), Controls: 0/391 (0%)<br>E388K: PD: 1/1130 (0.09%), Controls: 1/391 (0.26%)<br>D409H: PD: 1/1130 (0.09%), Controls: 0/391 (0%)<br>L444P: PD: 13/1130 (1.15%), Controls: 0/391 (0%) |

| | | | | | |
|---|---|---|---|---|---|
| | | | | | P452L: PD: 1/1130 (0.09%), Controls: 0/391 (0%)<br>R463C: PD: 1/1130 (0.09%), Controls: 0/391 (0%)<br>R463H: PD: 1/1130 (0.09%), Controls: 0/391 (0%)<br>G113A/A446A: PD: 1/1130 (0.09%), Controls: 0/391 (0%)<br>RecΔ5: PD: 2/1130 (0.18%), Controls: 0/391 (0%)<br>RecNciI (L444P+A456P+V460V): PD: 2/1130 (0.18%), Controls: 0/391 (0%)<br>RecA456P (L444P+A456P): PD: 1/1130 (0.09%), Controls: 0/391 (0%)<br>c.1263del+RecTL (c.1263–1317del+D409H+L444P+A456P+V460V): PD: 1/1130 (0.09%), Controls: 0/391 (0%)<br>D140H+E326K/E326K: PD: 1/1130 (0.09%), Controls: 0/391 (0%)<br>A190A: PD: 1/1130 (0.09%), Controls: 0/391 (0%)<br>Y313Y: PD: 0/1130 (0%), Controls: 1/391 (0.26%) |
| Spataro et al., 2017[16] (28124432) | 249/145 | Spanish | Targeted reseqeuncing and CNF detection by eXome- Hidden Markov Model (XHMM) software | NA | N370S | N370S: 1/249 (0.4%) |
| Setó-Salvia et al., 2012[17] (22173904) | 225/186 | Spanish | Cycle sequencing of *GBA* coding region | All *GBA* exons | N370S, L444P, L144V, S488T, M123T, G202R, I260T, T369M, W393R, D409H, RecNcil | N370S: PD: 5/225 (2.22%), Controls: 0/186 (0%)<br>L444P: PD: 6/225 (2.67%), Controls: 0/186 (0%)<br>L144V: PD: 1/225 (0.44%), Controls: 0/186 (0%)<br>S488T: PD: 1/225 (0.44%), Controls: 0/186 (0%)<br>M123T: PD: 1/225 (0.44%), Controls: 0/186 (0%)<br>G202R: PD: 1/225 (0.44%), Controls: 0/186 (0%)<br>I260T: PD: 1/225 (0.44%), Controls: 0/186 (0%)<br>T369M: PD: 2/225 (0.89%), Controls: 1/186 (0.5%)<br>W393R: PD: 1/225 (0.44%), Controls: 0/186 (0%)<br>D409H: PD: 2/225 (0.89%), Controls: 0/186 (0%)<br>RecNcil: PD: 1/225 (0.44%), Controls: 0/186 (0%) |
| Jesús et al., 2016[18] (28030538) | 532/542 | Southern Spanish | High-resolution melting (HRM) analysis and direct DNA resequencing | Full *GBA* gene | N370S, L444P, W312R, V457D, E326K, T369M, etc. | All variants: PD: (12.2%), Controls: (7.9%)<br>N370S: PD: 5/532 (0.94%)<br>L444P: PD: 13/532 (2.44%), Controls: 6/542 (1.11%)<br>W312R: PD: 6/532 (1.13%), Controls: 2/542 (0.37%)<br>V457D: PD: 3/532 (0.56%), Controls: 4/542 (0.74%)<br>c.116-8C>T: PD: 4/532 (0.75%), Controls: 7/542 (1.29%)<br>E326K: PD: 16/532 (3.00%), Controls: 13/542 (2.40%)<br>T369M: PD: 5/532 (0.94%), Controls: 2/542 (0.37%) |
| Bras et al., 2009[19] (18160183) | 230/430 | Portuguese | DNA sequencing of the complete open-reading frame, as well as intron/exon boundaries, of the *GBA* gene | All coding exons and exon/intron boundaries of the *GBA* gene | N409S, N435T, D448H, L483P, K13R, R41L, E365K, T408M, E427K | Pathogenic variants: PD: 14/230 (6.1%), Controls: 3/430 (0.7%)<br>N409S: PD: 5/230 (2.2%), Controls: 3/430 (0.7%)<br>N435T: PD: 5/230 (2.2%), Controls: 0/430 (0%)<br>D448H: PD: 1/230 (0.4%), Controls: 0/430 (0%)<br>L483P: PD: 3/230 (1.3%), Controls: 0/430 (0%)<br>K13R: PD: 1/230 (0.4%), Controls: 0/430 (0%)<br>R41L: PD: 0/230 (0%), Controls: 1/430 (0.2%)<br>E365K: PD: 2/230 (0.9%), Controls: 3/430 (0.7%)<br>T408M: PD: 2/230 (0.9%), Controls: 5/430 (1.2%)<br>E427K: PD: 0/230 (0%), Controls: 2/430 (0.5%) |
| Petrucci et al., 2020[20] (32658388) | 874/0 | Italian | Whole exome sequencing | All *GBA* exons | D24N, S107L, R120W, R131C, P182L, N188S, G202R, H255Q, D409H, L444P, R463C, W209Gfs*6, R257*, E388K, S196P, G202R, H255Q, D409H, T369M, L444P, A456P, V460V, G46E, G193R, R329C, N370S, E326K, N188K, W184R, I161N, K(-27)R, M85V, E326D, T369T, V460L | All: PD: 125/874 (14.3%)<br>N370S: PD: 30/874 (3.43%)<br>L444P: PD: 29/874 (3.32%)<br>E326K: PD: 16/874 (1.83%) |

| | | | | | | |
|---|---|---|---|---|---|---|
| De Marco et al., 2008[21] (18074383) | 395/483 | Italian | Genotyping | L444P, N370S | L444P, N370S | All: PD: 11/395 (2.8%), Controls: 1/483 (0.2%)<br>L444P: PD: 8/395 (2.0%), Controls: 1/483 (0.2%)<br>N370S: PD: 3/395 (0.8%), Controls: 0/483 (0%) |
| Asselta et al., 2014[22] (25249066) | 2350/1111 | Italian | High-resolution melting (HRM) analysis (exon 9) and direct DNA sequencing (exon 10) | *GBA* exons 9 and 10 | IVS8-24T>G, N370S, E388K, IVS9+32C>T, IVS9-36C>G, IVS9-5T>A, D443N, L444P, IVS10+1G>T, IVS10+8C>A | N370S or D443N or L444P or IVS10+1G>T: PD: 106/2350 (4.5%), Controls: 7/1111 (0.63%)<br>N370S: PD+DLB+MSA+PSP+CBD: 69/2766 (2.5%), Controls: 4/1111 (0.36%)<br>L444P: PD+DLB+MSA+PSP+CBD: 47/2766 (1.7%), Controls: 3/1111 (0.27%) |
| Cilia et al., 2016[23] (27632223) | 2843/0 | Italian | Variant screening of *GBA* exons 9 and 10 | *GBA* exons 9 and 10 | N370S, L444P, G377S, IVS10+1G>T | N370S: PD: 70/2843 (2.46%)<br>L444P: PD: 54/2843 (1.90%)<br>G377S: PD: 1/2843 (0.04%)<br>IVS10+1G>T: PD: 1/2843 (0.04%) |
| Straniero et al., 2020[24] (33209983) | 3691/7757 (1625 partners and caregivers of patients with PD) | Italian | Variant screening | E326K, T369M, N370S, L444P | E326K, T369M, N370S, L444P | E326K: PD: 61/3691 (1.65%), Controls: 55/7755 (0.71%)<br>T369M: PD: 49/3691 (1.33%), Controls: 61/7755 (0.79%)<br>N370S: PD: 76/3691 (2.06%), Controls: 43/7755 (0.55%)<br>L444P: PD: 62/3691 (1.68%), Controls: 11/7755 (0.14%) |
| Quadri et al., 2015[25] (25294124) | 100/0 | Sardinian | Whole exome sequencing | All *GBA* exons | N370S, R131C | N370S: PD: 4/100 (4%)<br>R131C: PD: 2/100 (2%) |
| Kalinderi et al., 2009[26] (19383421) | 172/132 | Greek | DNA sequencing of all *GBA* exons | All *GBA* exons | L445P, D409H, E326K, H255Q, R329H, L268L, S271G, T428K, V460L | L445P: PD: 2/172 (1.2%), Controls: 0/132 (0%)<br>D409H: 1/172 (0.6%), Controls: 0/132 (0%)<br>E326K: 1/172 (0.6%), Controls: 1/132 (0.8%)<br>H255Q: 4/172 (2.3%), Controls: 0/132 (0%)<br>R329H: 1/172 (0.6%), Controls: 0/132 (0%)<br>L268L+S271G: 1/172 (0.6%), Controls: 0/132 (0%)<br>T428K: 1/172 (0.6%), Controls: 1/132 (0.8%)<br>V460L: 0/172 (0%), Controls: 4/132 (3.0%) |
| Moraitou et al., 2011[27] (21745757) | 205/206 | Greek | Restriction enzyme analysis for eight variants | N370S, D409H, L444P, H255Q, R120W, Y108C, IVS10-1G>A, IVS6-2A>G | N370S, D409H, L444P, H255Q, Y108C, IVS10-1G>A, IVS6-2A>G | N370S: PD: 6/205 (2.93%), Controls: 4/206 (1.94%)<br>D409H: PD: 7/205 (3.41%), Controls: 0/206 (0%)<br>L444P: PD: 6/205 (2.93%), Controls: 1/206 (0.49%)<br>H255Q: PD: 7/205 (3.41%), Controls: 1/206 (0.49%)<br>Y108C: PD: 0/205 (0%), Controls: 1/206 (0.49%)<br>IVS10-1G>A: PD: 1/205 (0.49%), Controls: 0/206 (0%) |
| Emekli et al., 2021[28] (34781237) | 82/0 | Turkish | Next-generation sequencing | All GBA exons and intron/exon boundaries | R434P, H294Q, D448H, G241R, N227K | R434P: PD: 2/82 (2.4%)<br>H294Q: PD: 1/82 (1.2%)<br>D448H: PD: 1/82 (1.2%)<br>G241R: PD: 1/82 (1.2%)<br>N227K: PD: 1/82 (1.2%) |
| Kumar et al., 2013[29] (22812582) | 360/348 | Serbian | DNA sequencing of *GBA* exons 8-11 | *GBA* exons 8-11 | N370S, D409H, H255Q, L444P, A456P, R463C, RecNciI, T369M, E388K, D380V, N392S, V459V | All: PD: 21/360 (5.8%), Controls: 5/348 (1.4%)<br>N370S: PD: 9/360 (2.5%), Controls: 0/348 (0%)<br>D409H, H255Q: PD: 7/360 (1.9%), Controls: 2/348 (0.6%)<br>L444P: PD: 2/360 (0.6%), Controls: 1/348 (0.3%)<br>A456P: PD: 0/360 (0%), Controls: 1/348 (0.3%)<br>R463C: PD: 1/360 (0.3%), Controls: 0/348 (0%)<br>RecNciI (L444P+A456P+V460V): PD: 1/360 (0.3%), Controls: 0/348 (0%)<br>E388K: PD: 0/360 (0%), Controls: 1/348 (0.3%)<br>D380V: PD: 1/360 (0.3%), Controls: 0/348 (0%)<br>N329S: PD: 1/360 (0.3%), Controls: 0/348 (0%)<br>V459V: PD: 0/360 (0%), Controls: 1/348 (0.3%) |
| Török et al., 2016[30] (26547032) | 124/122 | Hungarian | Variant screening | L444P, N370S, R120W | L444P | L444P: PD: 3/124 (2.4%), Controls: 0/122 (0%) |
| Benitez et al., 2016[31] (27094865) | 478/337 | European-American | Deep-sequencing of all *GBA* exons | All *GBA* exons | R83C, H294Q, T336S, E365K, T408M, N409S, E427K, D448H, L483P, A495P | R83C: PD: 2/478 (0.41%), Controls: 0/337 (0%)<br>H294Q: PD: 2/478 (0.41%), Controls: 0/337 (0%)<br>T336S: PD: 1/478 (0.21%), Controls: 0/337 (0%)<br>E365K: PD: 19/478 (3.97%), Controls: 11/337 (3.26%) |

| | | | | | | |
|---|---|---|---|---|---|---|
| | | | | | | T408M: PD: 17/478 (3.56%), Controls: 0/337 (0%)<br>N409S: PD: 7/478 (1.46%), Controls: 1/337 (0.30%)<br>E427K: PD: 1/478 (0.21%), Controls: 0/337 (0%)<br>D448H: PD: 1/478 (0.21%), Controls: 1/337 (0.30%)<br>L483P: PD: 7/478 (1.46%), Controls: 2/337 (0.59%)<br>A495P: PD: 17/478 (3.56%), Controls: 10/337 (2.97%) |
| Noreau et al., 2011[32]<br>(21856586) | 212/189 | French-Canadian | Sequencing of the entire coding region of *GBA* | All *GBA* exons | L197F, E326K, S339L, T369M, N370S, W378G, L444P | L197F: PD: 1/212 (0.47%), Controls: 0/189 (0%)<br>E326K: PD: 5/212 (2.36%), Controls: 3/189 (1.59%)<br>S339L: PD: 1/212 (0.47%), Controls: 0/189 (0%)<br>T369M: PD: 9/212 (4.25%), Controls: 5/189 (2.65%)<br>N370S: PD: 0/212 (0%), Controls: 2/189 (1.06%)<br>W378G: PD: 1/212 (0.47%), Controls: 0/189 (0%)<br>L444P: PD: 5/212 (2.36%), Controls: 1/189 (0.53%) |
| Han et al., 2016[33]<br>(26000814) | 225/110<br>(spouses) | Canadian | DNA sequencing of full *GBA* gene | All *GBA* exons and flanking introns | c.-119A/G, S(-35)N, R120W, N370S, L444P, RecNciI, RecTL (del55/D409H/RecNciI), E326K, T369M, S13L | All: PD: 25/225 (11.11%), Controls: 9/110 (8.19%)<br>c.-119A/G: PD: 1/225 (0.44%), Controls: 0/110 (0%)<br>S(-35)N: PD: 1/225 (0.44%), Controls: 0/110 (0%)<br>R120W: PD: 1/225 (0.44%), Controls: 0/110 (0%)<br>N370S: PD: 2/225 (0.89%), Controls: 0/110 (0%)<br>L444P: PD: 4/225 (1.78%), Controls: 0/110 (0%)<br>RecNciI (L444P-A456P-V460V): PD: 1/225 (0.44%), Controls: 0/110 (0%)<br>RecTL (del55/D409H/RecNciI): PD: 2/225 (0.89%), Controls: 0/110 (0%)<br>E326K: PD: 4/225 (1.78%), Controls: 4/110 (3.64%)<br>T369M: PD: 11/225 (4.89%), Controls: 4/110 (3.64%)<br>S13L: PD: 0/225 (0%), Controls: 1/110 (0.91%) |
| Sato et al., 2005[34]<br>(15517592) | 88/122 | Canadian | Genotyping | N370S, L444P, IVS2+1, K198T, R329C, 84insGG, Rec | N370S, L444P, Rec | N370S: PD: 1/88 (1.14%), Controls: 1/122 (0.82%)<br>L444P: PD: 1/88 (1.14%), Controls: 0/122 (0%)<br>Rec: PD: 3/88 (3.41%), Controls: 0/122 (0.%) |
| González-Del Rincón et al., 2013[35]<br>(23448517) | 128/252 (128 sex and age matched, 124 (aged >60)) | Mexican Mestizo | Variant screening | N370S, L444P | L444P | L444P: PD: 7/128 (5.47%), Controls: 0/252 (0%) |
| Tipton et al., 2020[36]<br>(32197197) | 209/58 | Colombian and Hispanic American | Variant screening | K198E | K198E | Colombian:<br>K198E: PD: 3 (2.1%), Controls: 1 (1.7%)<br>Hispanic American:<br>K198E: PD: 0 (0%) |
| Velez-Pardo et al., 2019[37]<br>(30765263) | 602/319 | Colombian, Peruvian | DNA sequencing of all *GBA* exons and intron/exon boundaries | All *GBA* exons and intron/exon boundaries | R86X, R159W, R170C, G234W, K237E, N409S, L483P, L483P + RecG or L483P + Rec6b (L483P, +92G>A), Rec1 (L483P, A495P, V499V), RecD, E, or AZRecTL (L483P, A495P, V499V, +92G>A), D66H, R250K, R316H, M400I, E427K, D482N, I528V, R534H, K13R, E365K, T408M | Colombian:<br>R86X: PD: 0/131 (0%), Controls: 0/164 (0%)<br>R159W: PD: 0/131 (0%), Controls: 0/164 (0%)<br>R170C: PD: 0/131 (0%), Controls: 0/164 (0%)<br>G234W: PD: 0/131 (0%), Controls: 1/164 (0.6%)<br>K237E: PD: 7/131 (5.3%), Controls: 2/164 (1.2%)<br>N409S: PD: 3/131 (2.3%), Controls: 0/164 (0%)<br>L483P: PD: 3/131 (2.3%), Controls: 0/164 (0%)<br>L483P + RecG or L483P + Rec6b (L483P, +92G>A): PD: 0/131 (0%), Controls: 0/164 (0%)<br>Rec1 (L483P, A495P, V499V): PD: 0/131 (0%), Controls: 0/164 (0%)<br>RecD, E, or AZRecTL (L483P, A495P, V499V, +92G>A): PD: 0/131 (0%), Controls: 0/164 (0%)<br>D66H: PD: 0/131 (0%), Controls: 0/164 (0%)<br>R250K: PD: 0/131 (0%), Controls: 1/164 (0.6%)<br>R316H: PD: 0/131 (0%), Controls: 0/164 (0%)<br>M400I: PD: 0/131 (0%), Controls: 0/164 (0%)<br>E427K: PD: 0/131 (0%), Controls: 1/164 (0.6%)<br>D482N: PD: 0/131 (0%), Controls: 0/164 (0%)<br>I528V: PD: 0/131 (0%), Controls: 0/164 (0%)<br>R534H: PD: 0/131 (0%), Controls: 1/164 (0.6%) |

| | | | | | | |
|---|---|---|---|---|---|---|
| | | | | | | K13R: PD: 1/131 (0.8%), Controls: 1/164 (0.6%)<br>E365K: PD: 2/131 (1.5%), Controls: 1/164 (0.6%)<br>T408M: PD: 0/131 (0%), Controls: 0/164 (0%)<br><br>Peruvian:<br>R86X: PD: 1/471 (0.2%), Controls: 0/155 (0%)<br>R159W: PD: 2/471 (0.4%), Controls: 0/155 (0%)<br>R170C: PD: 3/471 (0.6%), Controls: 0/155 (0%)<br>G234W: PD: 0/471 (0%), Controls: 0/155 (0%)<br>K237E: PD: 0/471 (0%), Controls: 0/155 (0%)<br>N409S: PD: 1/471 (0.2%), Controls: 1/155 (0.6%)<br>L483P: PD: 7/471 (1.5%), Controls: 0/155 (0%)<br>L483P + RecG or L483P + Rec6b (L483P, +92G>A): PD: 1/471 (0.2%), Controls: 0/155 (0%)<br>Rec1 (L483P, A495P, V499V): PD: 4/471 (0.8%), Controls: 1/155 (0.6%)<br>RecD, E, or AZRecTL (L483P, A495P, V499V, +92G>A): PD: 1/471 (0.2%), Controls: 0/155 (0%)<br>D66H: PD: 1/471 (0.2%), Controls: 0/155 (0%)<br>R250K: PD: 0/471 (0%), Controls: 0/155 (0%)<br>R316H: PD: 1/471 (0.2%), Controls: 0/155 (0%)<br>M400I: PD: 0/471 (0%), Controls: 1/155 (0.6%)<br>E427K: PD: 0/471 (0%), Controls: 0/155 (0%)<br>D482N: PD: 1/471 (0.2%), Controls: 0/155 (0%)<br>I528V: PD: 1/471 (0.2%), Controls: 0/155 (0%)<br>R534H: PD: 0/471 (0%), Controls: 0/155 (0%)<br>K13R: PD: 3/471 (0.6%), Controls: 0/155 (0%)<br>E365K: PD: 5/471 (1.1%), Controls: 0/155 (0%)<br>T408M: PD: 3/471 (0.6%), Controls: 0/155 (0%) |
| Eblan et al., 2006[38]<br>(16261622) | 33/31 | Venezuelan | DNA sequencing of all *GBA* exons and most flanking introns | All *GBA* exons and most flanking introns | N370S, L444P, RecNciI, D443N | N370S: PD: 1/33 (3.0%), Controls: 0/31 (0%)<br>L444P: PD: 1/33 (3.0%), Controls: 0/31 (0%)<br>RecNciI: PD: 2/33 (6.1%), Controls: 0/31 (0%)<br>D443N: PD: 0/33 (0%), Controls: 1/31 (3.32%) |
| Dos Santos et al., 2010[39]<br>(20816920) | 110/155 | Brazilian | Variant screening | N370S, L444P, 84GG, IVS2+1G>A, G377S | N370S, L444P, D409H+L444P+A456P+V460V, IVS2+1G>A | N370S: PD: 2/110 (1.8%), Controls: 0/155 (0%)<br>L444P: PD: 2/110 (1.8%), Controls: 0/155 (0%)<br>D409H+L444P+A456P+V460V: PD: 1/110 (0.9%), Controls: 0/155 (0%)<br>IVS2+1G>A: PD: 1/110 (0.9%), Controls: 0/155 (0%) |
| Spitz et al., 2008[40]<br>(17703984) | 65/267 | Brazilian | Variant screening | L444P, N370S | L444P | L444P: PD: 2/65 (3.1%), Controls: 0/267 (0%) |
| Guimarães Bde et al., 2012[41]<br>(22192918) | 237/186 | Brazilian | Direct sequencing | N370S, L444P | N370S, L444P | L444P: PD: 3/237 (1.27%)<br>N370S: PD: 6/237 (2.53%) |
| Socal et al., 2009[42]<br>(18358758) | 62/0 | Brazilian (with mixed ethnic backgrounds) | Variant screening | L444P, N370S, IVS2+1, 84GG | L444P, N370S | L444P: PD: 1/62 (1.61%)<br>N370S: PD: 1/62 (1.61%) |
| Barkhuizen et al., 2017[43]<br>(28361101) | 105/40 | Caucasian/South African (82.7% Afrikaner) | Sanger sequencing of *GBA* exons 8-11 in all participants; direct sequencing of all *GBA* exons in 20 PD cases | All *GBA* exons; *GBA* exons 8-11 | G35A, E326K, I368T, T369M, N370S, P387L, K441N | G35A: PD: 1/20 (5.0 %)<br>E326K: PD: 5/105 (4.8%), Controls: 1/40 (2.5%)<br>I368T: PD: 1/105 (1.0%), Controls: 0/40 (0%)<br>T369M: PD: 2/105 (1.9%), Controls: 1/40 (2.5%)<br>N370S: PD: 1/105 (1.0%), Controls: 0/40 (0%)<br>P387L: PD: 2/105 (1.9%), Controls: 0/40 (0%)<br>K441N: PD: 1/105 (1.0%), Controls: 0/40 (0%) |
| Mahungu et al., 2020[44]<br>(32035846) | 30/0 | Black South African | Sanger sequencing of all *GBA* exons | All *GBA* exons | K13R, T75del, R159W, R170L, F255L, Q536, G517R, Q471Q | K13R: PD: 6/30 (20%)<br>T75del: PD 2/30 (6.6%)<br>R159W: PD: 1/30 (3.3%)<br>R170L: PD: 1/30 (3.3%)<br>F255L: PD: 6/30 (6.6%)<br>Q536: PD 1/30 (together with F255L) (3.3%) |

| | | | | | | |
|---|---|---|---|---|---|---|
| | | | | | | G517R: PD 1/30 (3.3%)<br>Q471Q: PD: 1/30 (3.3%) |
| Lesage et al., 2011[45]<br>(21242499) | 194/177 | North African (PD: Algeria: n=147, Morocco: n=23, Tunisia: n=14, Libya: n=1, unknown: n=9; Controls: Algeria: n=95, Morocco: n=46) | DNA sequencing of *GBA* coding regions | All *GBA* exons | K(-27)R, R131C, N370S, L444P, E326K, RecNciI (A456P/V460V/L444P), D443N, T369M | K(-27)R: PD: 2/194 (1.03%), Controls: 0/177 (0%)<br>R131C: PD: 2/194 (1.03%), Controls: 0/177 (0%)<br>N370S: PD: 2/194 (1.03%), Controls: 0/177 (0%)<br>L444P/E326K: 1/194 (0.52%), Controls: 0/177 (0%)<br>RecNciI (A456P/V460V/L444P): PD: 2/194 (1.03%), Controls: 0/177 (0%)<br>D443N: PD: 0/194 (0%), Controls: 1/177 (0.56%)<br>E326K: PD: 1/194 (0.52%), Controls: 0/177 (0%)<br>T369M: PD: 2/194 (1.03%), Controls: 0/177 (0%) |
| Nishioka et al., 2010[46]<br>(19945510) | 395/372 | North African Arab-Berber | DNA sequencing of all *GBA* exons | All *GBA* exons | K13R, K225R, N370S | K13R: PD: 2/33 (3.0%)<br>K225R: PD: 1/33 (6.1%)<br>K13R: PD: 5/155 (familial, including the 33) (3.2%), 9/240 (sporadic) (3.8%), Controls: 16/372 (4.3%)<br>K225R: PD: 2/155 (familial, including the 33) (1.3%), 0/240 (sporadic) (0%), Controls: 0/372 (0%)<br>N370S: PD: 0/155 (familial, including the 33) (0%), 1/240 (sporadic) (0.4%), Controls: 3/372 (0.8%) |
| Emelyanov et al., 2018[47]<br>(30146349) | 762/400 | Russian | Variant screening | L444P, N370S, E326K, T369M | L444P, N370S, E326K, T369M | L444P: PD: 9/762 (1.1%), Controls: 1/400 (0.1%)<br>N370S: PD: 4/762 (0.5%), Controls: 0/400 (0%)<br>E326K: PD: 14/762 (2.4%), Controls: 5/400 (1.3%)<br>T369M: PD: 15/762 (2.5%), Controls: 4/400 (1.1%) |
| Emelyanov et al., 2012[48]<br>(21915911) | 330/240 | Russian | Variant screening | L444P, N370S | L444P, N370S | L444P: PD: 6/330 (1.8%), Controls: 1/240 (0.5%)<br>N370S: PD: 3/330 (0.9%), Controls: 0/240 (0%) |
| Mao et al., 2010[49]<br>(20004703) | 616/411 | Han-Chinese | Variant screening | L444P | L444P | L444P: PD: 20/616 (3.2%), Controls: 1/411 (0.2%) |
| Hu et al., 2010[50]<br>(20528910) | 328/300 | Han Chinese | Genotyping | N370S | N370S | N370S: PD: 6/328 (1.8%), Controls: 2/300 (0.7%) |
| Zhang et al., 2012[51]<br>(23286447) | 195/443 | Han Chinese | Genotyping | L444P, N370S, R120W | L444P | L444P: PD: 6/195 (3.08%), Controls: 0/443 (0%) |
| Guo et al., 2015[52]<br>(25623333) | 1019/1030 | Han-Chinese | Genotyping | L444P | L444P | L444P: PD 26/1019 (2.7%), Controls: 1/1030 (0.1%) |
| Wang et al., 2014[53]<br>(24095219) | 1638/0 | Han Chinese | Genotyping | L444P | L444P | L444P: PD 49/1638 (2.99%) |
| Ren et al., 2022[54]<br>(34951095) | 737/0 | Chinese | Next-generation sequencing | Full *GBA* gene | N370S, E326K, T369M, R163Q, L444P, R120W, etc. | All variants: PD: 79/737 (10.72%)<br>Mild (e.g. N370S): PD: 8/79 (10.13%) -> 8/737 (1.09%)<br>Severe (e.g. L444P): PD: 28/79 (35.44%) -> 28/737 (3.80%)<br>Risk (e.g. E326K): PD: 1/79 (1.27%) -> 1/737 (0.14%)<br>Complex (e.g. L444P-A456P-V460V): PD: 7/79 (8.86%) -> 7/737 (0.95%)<br>Unknown: PD: 35/79 (44.3%) -> 35/737 (4.75%)<br><br>R163Q: PD: 12/79 (15.19%) -> 12/737 (1.63%)<br>L444P: PD: 10/79 (12.66%) -> 10/737 (1.36%)<br>R120W: PD: 6/79 (7.59%) -> 6/737 (0.81%) |
| Yu et al., 2015[55]<br>(25518742) | 184/130 | Chinese | DNA sequencing of all *GBA* exons | All *GBA* exons | R163Q, F213I, E326K, S364S, F347L, V375L, L444P, RecNciI (L444P-A456P-V460V), A456P Q497R, c.334_338delCAGAA, L264I, L314V | All: PD: 16/184 (8.7%), Controls: 2/130 (1.54%)<br>R163Q: PD: 1/184 (0.54%), Controls: 0/130 (0%)<br>F213I: PD: 1/184 (0.54%), Controls: 0/130 (0%)<br>E326K: PD: 1/184 (0.54%), Controls: 0/130 (0%)<br>S364S: PD: 1/184 (0.54%), Controls: 0/130 (0%)<br>F347L: PD: 1/184 (0.54%), Controls: 1/130 (0.77%)<br>V375L: PD: 1/184 (0.54%), Controls: 0/130 (0%)<br>L444P: PD: 2/184 (1.09%), Controls: 0/130 (0%)<br>RecNciI (L444P-A456P-V460V): PD: 3/184 (1.63%), Controls: 0/130 (0%)<br>A456P: PD: 0/184 (0%), Controls: 1/130 (0.77%) |

| | | | | | | |
|---|---|---|---|---|---|---|
| | | | | | | Q497R: PD: 1/184 (0.54%), Controls: 0/130 (0%)<br>c.334_338delCAGAA: PD: 1/184 (0.54%), Controls: 0/130 (0%)<br>L264I: PD: 2/184 (1.09%), Controls: 0/130 (0%)<br>L314V: PD: 1/184 (0.54%), Controls: 0/130 (0%) |
| Sun et al., 2010[56] (20131388) | 402/413 | Chinese | Variant screening | L444P, F213I, R353W, N370S | L444P | L444P: PD: 11/402 (2.74%), Controls: 0/413 (0%) |
| Wang et al., 2012[57] (23227814) | 208/298 | Chinese | Variant screening | L444P, N370S, R120W | L444P | L444P: PD: 7/208 (3.4%), Controls: 1/298 (0.3%)<br>N370S: PD: 0/208 (0%), Controls: 0/298 (0%)<br>R120W: PD: 0/208 (0%), Controls: 0/298 (0%) |
| Ziegler et al., 2007[58] (17462935) | 92/92 | Chinese | Direct sequencing of full *GBA* gene | All *GBA* exons and flanking introns | L444P, D409H, L174P, Q497R, V460M | L444P: PD: 1/92 (1.1%), Controls: 0/92 (0%)<br>D409H: PD: 1/92 (1.1%), Controls: 0/92 (0%)<br>L174P: PD: 1/92 (1.1%), Controls: 0/92 (0%)<br>Q497R: PD: 1/92 (1.1%), Controls: 0/92 (0%)<br>V460M: PD: 0/92 (0%), Controls: 1/92 (1.1%) |
| Tan et al., 2007[59] (17620502) | 331/347 | Chinese | Allelic discrimination using the 5' nuclease activity assay, adapted to detect the L444P and N370S variants | L444P, N370S | L444P | L444P: PD: 8/331 (2.4%), Controls: 0/347 (0%) |
| Li et al., 2020[60] (32171587) | 240/0 | Chinese | Whole-exome sequencing | All *GBA* exons | IVS2+1, G202R, D409H, L444P | IVS2+1: PD: 1/240 (0.4%)<br>G202R: PD: 1/240 (0.4%)<br>D409H: PD: 2/240 (0.8%)<br>L444P: PD: 1/240 (0.4%) |
| Huang et al., 2011[61] (21338444) | 967/780 (spouses, patients with unrelated diseases, healthy volunteers) | Chinese | DNA sequencing of whole *GBA* coding region (in 30 PD cases); Genotyping of L444P, D409H, R120W, L174P, Q497R in all participants | All *GBA* exons; L444P, D409H, R120W, L174P, Q497R | L444P, D409H, RecNciI (L444P-A456P-V460V) | All: PD: 36/967 (3.72%), Controls: 2/780 (0.26%)<br>L444P: PD: 27/967 (2.79%), Controls: 1/780 (0.13%)<br>D409H: PD: 2/967 (0.21%), Controls: 0/780 (0%)<br>RecNciI (L444P-A456P-V460V): PD: 7/967 (0.72%), Controls: 1/780 (0.13%) |
| Zhang et al., 2015[62] (26421210) | 1147/0 | Chinese | Variant screening | L444P | L444P | L444P: PD: 34/1147 (2.96%) |
| Foo et al., 2014[63] (24565865) | 1085/9445 | Chinese, Korean | Sequencing of all *GBA* exons in EOPD cases and matching controls; genotyping in LOPD cases and matching controls | All *GBA* exons | Various, including S350R, V221I, P210fs, c.281+1G>A | Chinese EOPD: All rare and low frequency variants: PD: 4/195 (2.05%), Controls: 3/219 (1.37%)<br>Korean EOPD: All rare and low frequency variants: PD: 18/180 (10.0%), Controls: 6/180 (3.33%)<br><br>Chinese LOPD: All rare and low frequency variants: PD: 0/710 (0%), Controls: 3/9046 (0.03%) |
| Gutti et al., 2008[64] (18541817) | 184/0 | Chinese from Taiwan | Sequencing of *GBA* gene | Full *GBA* gene | L444P, R131S, R163Q, L174P, S271G, D409H, Q497R | L444P: PD: 4/184 (2.17%)<br>R131S: PD: 1/184 (0.54%)<br>R163Q: PD: 1/184 (0.54%)<br>L174P: PD: 1/184 (0.54%)<br>S271G: PD: 1/184 (0.54%)<br>D409H: PD: 1/184 (0.54%)<br>Q497R: PD: 1/184 (0.54%) |
| Wu et al., 2007[65] (17702778) | 518/339 | Taiwanese | Variant screening | L444P, RecNciI, R120W | L444P, RecNciI, R120W | L444P: PD: 13/518 (2.5%), Controls: 2/339 (0.6%)<br>RecNciI: PD: 2/518 (0.4%), Controls: 2/339 (0.6%)<br>R120W: PD: 1/518 (0.2%), Controls: 0/339 (0%) |

| | | | | | | |
|---|---|---|---|---|---|---|
| Choi et al., 2012[66] (22387070) | 277/291 | Korean | Direct DNA sequencing of all *GBA* exons in 277 PD cases and 100 controls, only exon 2 and exons 5–11 in 191 controls | All *GBA* exons; *GBA* exons 2, 5-11 | I-20V, R163Q, N188S, P201H, R257Q, L268L, S271G, R277C, F347L, L444P, K466K | I-20V: PD: 1/277 (0.36%), Controls: 4/291 (1.37%)<br>R163Q: PD: 1/277 (0.36%), Controls: 0/291 (0%)<br>N188S: PD: 1/277 (0.36%), Controls: 0/291 (0%)<br>P201H: PD: 1/277 (0.36%), Controls: 0/291 (0%)<br>R257Q: PD: 3/277 (1.08%), Controls: 0/291 (0%)<br>L268L: PD: 1/277 (10.36%), Controls: 0/291 (0%)<br>S271G: PD: 2/277 (0.72%), Controls: 0/291 (0%)<br>R277C: PD: 1/277 (0.36%), Controls: 0/291 (0%)<br>F347L: PD: 1/277 (0.36%), Controls: 0/291 (0%)<br>L444P: PD: 2/277 (0.72%), Controls: 0/291 (0%)<br>K466K: PD: 4/277 (1.44%), Controls: 1/291 (0.34%) |
| Li et al., 2014[67] (24126159) | 147/100 | Japanese | DNA sequencing of all *GBA* exons and intron/exon boundaries | All *GBA* exons and exon/intron boundaries of *GBA* | I(-20)V, G64V, R120W, D409H, L444P, I489V, W393X, K466K, c.1447-1466delTGins, RecNciI | I(-20)V: PD: 13/144 (9.0%), Controls: 10/100 (10.0%)<br>G64V: PD: 1/144 (0.7%), Controls: 0/100 (0%)<br>R120W: PD: 9/144 (6.3%), Controls: 0/100 (0%)<br>D409H: PD: 4/144 (2.8%), Controls: 0/100 (0%)<br>L444P: PD: 12/144 (8.3%), Controls: 0/100 (0%)<br>I489V: PD: 2/144 (1.4%), Controls: 0/100 (0%)<br>W393X: PD: 1/144 (0.7%), Controls: 0/100 (0%)<br>c.1447-1466delTGins: PD: 1/144 (0.7%), Controls: 0/100 (0%)<br>RecNciI: PD: 1/144 (0.7%), Controls: 1/100 (1.0%) |
| Mitsui et al., 2009[68] (19433656) | 534/544 | Japanese | DNA sequencing of all *GBA* exons | All *GBA* exons | R120W, R131C, N188S, R120W-N188R-V191G-S196P-F213I, G193W, F213I, R329C, L444P, L444P-A456P-V460V (RecNciI), A456P-V460V, R496C, I(-20), L(-15)F, L67Q, V121V, D153N, R163Q, P299T, G307S, T334I, L336L, G344G, F347L, R359L, V460V, K466K, I489V | R120W: PD: 15/534 (2.8%), Controls: 0/544 (0%)<br>R131C: PD: 1/534 (0.2%), Controls: 0/544 (0%)<br>N188S: PD: 4/534 (0.7%), Controls: 0/544 (0%)<br>R120W-N188R-V191G-S196P-F213I: PD: 1/534 (0.2%), Controls: 0/544 (0%)<br>G193W: PD: 1/534 (0.2%), Controls: 0/544 (0%)<br>F213I: PD: 1/534 (0.2%), Controls: 0/544 (0%)<br>R329C: PD: 2/534 (0.4%), Controls: 0/544 (0%)<br>L444P: PD: 8/534 (1.5%), Controls: 0/544 (0%)<br>L444P-A456P-V460V (RecNciI): PD: 14/534 (2.6%), Controls: 2/544 (0.4%)<br>A456P-V460V: PD: 1/534 (0.2%), Controls: 0/544 (0%)<br>R496C: PD: 2/534 (0.4%), Controls: 0/544 (0%)<br>I(-20)V: PD: 77/534 (14.4%), Controls: 66/544 (12.1%)<br>L(-15)F: PD: 1/534 (0.2%), Controls: 0/544 (0%)<br>L67Q: PD: 0/534 (0%), Controls: 1/544 (0.2%)<br>V121V: PD: 0/534 (0%), Controls: 1/544 (0.2%)<br>D153N: PD: 1/534 (0.2%), Controls: 0/544 (0%)<br>R163Q: PD: 4/534 (0.7%), Controls: 7/544 (1.3%)<br>P299T: PD: 1/534 (0.2%), Controls: 0/544 (0%)<br>G307S: PD: 1/534 (0.2%), Controls: 0/544 (0%)<br>T334I: PD: 0/534 (0%), Controls: 1/544 (0.2%)<br>L336L: PD: 1/534 (0.2%), Controls: 0/544 (0%)<br>G344G: PD: 1/534 (0.2%), Controls: 0/544 (0%)<br>F347L: PD: 0/534 (0%), Controls: 1/544 (0.2%)<br>R359L: PD: 1/534 (0.2%), Controls: 0/544 (0%)<br>V460V: PD: 2/534 (0.4%), Controls: 1/544 (0.2%)<br>K466K: PD: 11/534 (2.1%), Controls: 8/544 (1.5%)<br>I489V: PD: 4/534 (0.7%), Controls: 3/544 (0.6%) |
| Pulkes et al., 2014[69] (24997549) | 480/395 | Thai | Direct DNA sequencing in all EOPD and 100 patients with AAO>50; Variant screening in remaining patients with AAO>50 and controls | All *GBA* exons and exon/intron boundaries of *GBA* | L444P, N386K, P428S, IVS2+1G>A, IVS9+3G>C, IVS10-9_10GT>AG, V398fsX404 | L444P: PD: 15/480 (3.1%), EOPD: 8/108 (7.4%), AAO>50: 7/372 (1.9%), Controls: 1/395 (0.3%)<br>N386K: PD: 1/480 (0.2%), EOPD: 1/108 (0.9%), AAO>50: 0/372 (0%), Controls: 0/395 (0%)<br>P428S: PD: 2/480 (0.4%), EOPD: 1/108 (0.9%), AAO>50: 1/372 (0.3%), Controls: 1/395 (0.3%)<br>IVS2+1G>A: PD: 1/480 (0.2%), EOPD: 1/108 (0.9%), AAO>50: 0/372 (0%), Controls: 0/395 (0%)<br>IVS9+3G>C: PD: 1/480 (0.2%), EOPD: 1/108 (0.9%), AAO>50: 0/372 (0%), Controls: 0/395 (0%) |

| | | | | | | |
|---|---|---|---|---|---|---|
| | | | | | | IVS10-9_10GT>AG: PD: 3/480 (0.6%), EOPD: 1/108 (0.9%), AAO>50: 2/372 (0.5%), Controls: 0/395 (0%)<br>V398fsX404: PD: 1/480 (0.2%), EOPD: 1/108 (0.9%), AAO>50: 0/372 (0%), Controls: 0/395 (0%) |
| Yadav et al., 2018[70] (30504558) | 100/0 | Indian | DNA sequencing of all *GBA* exons and intron/exon boundaries | All *GBA* exons and exon/intron junctions | IVS1+191G>C, IVS4+47G>A, IVS6−86A>G, IVS9+141A>G, IVS10+3G>A | IVS1+191G>C: PD 1/100 (1%)<br>IVS4+47G>A: PD 64/100 (64%)<br>IVS6−86A>G: PD 65/100 (65%)<br>IVS9+141A>G: PD 65/100 (65%)<br>IVS10+3G>A: PD 1/100 (1%) |
| Biswas et al., 2021[71] (33711404) | 198/241 | Indian | Variant screening | IVS2+1A>G, R120W, H255Q, R257Q, E326K, N370S, D409H, L444P, RecNciI | L444P | L444P: PD: 2/198 (1.01%), Controls: 0/241 (0%) |
| Halder et al., 2016[72] (NA) | 114/120 | Indian | Variant screening | L444P, N370S | L444P | L444P: PD: 4/114 (3.51%), Controls: 0/120 (0%) |
| Goldstein et al., 2019[73] (31662221) | 1200/378 | Ashkenazi Jewish | Genotyping | E326K, T369M, R44C, N370S, R496H, L444P, 84GG, IVS2+1G->A, V394L, Rec370 | E326K, T369M, R44C, N370S, R496H, L444P, 84GG, IVS2+1G->A, V394L, Rec370 | E326K: PD: 17/1200 (1.4%), Controls: 5/378 (1.32%)<br>R44C: PD: 1/1200 (0.08%), Controls: 0/378 (0%)<br>T369M: PD: 11/1200 (0.92%), Controls: 1/378 (0.26%)<br>N370S or R535H: PD: 140/1200 (11.67%), Controls: 14/378 (3.7%)<br>84GG or IVS2+1 or V349L or L444P or Rec370: PD: 46/1200 (3.83%), Controls: 2/378 (0.53%) |
| Gan-Or et al., 2008[74] (18434642) | 420/4138 | Ashkenazi Jewish | Variant screening | N370S, R496H, 84GG IVS2+1, V394L, D409H, L444P, RecTL | N370S, R496H, 84GG, IVS2+1, V394L, D409H, L444P, RecTL | All: PD: 75/420 (17.9%), Elderly controls: (4.2%), Young controls: (6.35%)<br>N370S: PD: 46/420 (10.95%), Elderly controls: 11/333 (3.3%), Young controls: 224/3805 (5.89%)<br>R496H: PD: 7/420 (1.67%), Elderly controls: 1/333 (0.3%)<br>84GG: PD: 8/420 (1.90%), Elderly controls: 1/333 (0.3%), Young controls: 6/3805 (0.16%)<br>IVS2+1: PD: 4/420 (0.95%), Young controls: 4/3805 (0.11%)<br>V394L: PD: 3/420 (0.71%), Young controls: 4/3805 (0.11%)<br>L444P: PD: 2/420 (0.48%), Young controls: 2/3805 (0.05%) |
| Gan-Or et al., 2015[75] (25653295) | 1000/3805 | Ashkenazi Jewish | Variant screening | 84GG, IVS2+1, N370S, L444P, V394L, R496H, 370Rec | 84GG, IVS2+1, N370S, L444P, V394L, R496H, 370Rec | All: PD: 192/1000 (19.2%), Controls: 242/3805 (6.4%)<br>N370S: PD: 131/1000 (13.1%), Controls: 225/3805 (5.9%)<br>R496H: PD: 19/1000 (1.9%), Controls: NT<br>84GG: PD: 21/1000 (2.1%), Controls: 6/3805 (0.16%)<br>IVS2+1G>A: PD: 5/1000 (0.5%), Controls: 1/3805 (0.03%)<br>V394L: PD: 11/1000 (1.1%), Controls: 4/3805 (0.11%)<br>L444P: PD: 3/1000 (0.3%), Controls: 4/3805 (0.11%)<br>370Rec: PD: 10/1000 (1.0%), Controls: 2/3805 (0.05%) |
| Dagan et al., 2015[76] (26169695) | 287/400 | Ashkenazi Jewish | Variant screening | N370S, L444P, c.84GG, c.115+1G>A (IVS2+1G>A), V394L, R496H | N370S, c.84GG, V394L, R496H | N370S: PD: 54/287 (18.8%), Controls: 14/400 (3.5%)<br>c.84GG: PD: 9/287 (3.1%), Controls: 2/400 (0.5%)<br>V394L: PD: 1/287 (0.3%)<br>R496H: PD: 4/287 (1.4%), Controls: 3/400 (0.7%) |
| Liu et al., 2011[77] (21812969) | 268/178 | Ashkenazi Jewish | Genotyping | NA | N370S | N370S: PD: 28/268 (10.4%) |
| Ruskey et al., 2019[78] (29842932) | 735/622 | Ashkenazi Jewish | Targeted next-generation sequencing; Sanger sequencing of exons 10 and 11 | Full *GBA* gene | 84GG, R44C, N188S, E326K, T369M, N370S, A384D, V394L, T410M, L444P, L461P, R496H | 84GG: PD: 13/735 (1.77%), Controls: 1/622 (0.15%)<br>R44C: PD: 2/735 (0.27%), Controls: 7/622 (1.06%)<br>N188S: PD: 1/735 (0.14%), Controls: 0/622 (0%)<br>E326K: PD: 13/735 (1.77%), Controls: 2/622 (0.3%)<br>T369M: PD: 2/735 (0.27%), Controls: 0/622 (0%)<br>N370S: PD: 92/735 (12.52%), Controls: 37/622 (5.58%)<br>A384D: PD: 1/735 (0.14%), Controls: 0/622 (0%)<br>V394L: PD: 1/735 (0.14%), Controls: 0/622 (0%)<br>T410M: PD: 0/735 (0%), Controls: 1/622 (0.15%)<br>L444P: PD: 3/735 (0.41%), Controls: 0/622 (0%) |

| | | | | | | |
|---|---|---|---|---|---|---|
| | | | | | | L461P: PD: 1/735 (0.14%), Controls: 0/622 (0%)<br>R496H: PD: 9/735 (1.22%), Controls: 2/622 (0.3%) |
| Clark et al., 2005[79] (15517591) | 160/92 | Ashkenazi Jewish | Direct sequencing | N370S | N370S | N370S: PD: 17/160 (10.6%), Controls: 4/92 (4.3%) |
| Aharon-Peretz et al., 2004[80] (15525722) | 99/1543 | Ashkenazi Jewish | Variant screening | N370S, L444P, 84GG, IVS+1, V394L, R496H | N370S, 84GG, R496H | N370S: PD: 26/99 (26.3%), Controls: 92/1543 (5.96%)<br>84GG: PD: 4/99 (4.0%), Controls: 3/1543 (0.19%)<br>R496H: PD: 1/99 (1.0%), Controls: 0/1543 (0%) |
| Aharon-Peretz et al., 2005[81] (16148263) | 148/0 | Ashkenazi Jewish | Digestion with appropriate enzymes to detect N370S, L444P, 84GG, IVS+1, V394L, R496H | N370S, L444P, 84GG, IVS+1, V394L, R496H | N370S,4GG, R496H | N370S: PD: 34/148 (22.97%)<br>84GG: PD: 4/148 (2.70%)<br>R496H: 2/148 (1.35%) |
| Gan-Or et al., 2010[82] (19458969) | 600/0 | Ashkenazi Jewish | Variant screening | 84GG, IVS2+1, N370S, V394L, D409H, L444P, R496H and RecTL | 84GG, IVS2+1, N370S, V394L, D409H, L444P, R496H and RecTL | All: PD: 117/600 (19.5%) |
| Alcalay et al., 2015[83] (26117366) | 517/252 (mostly spouses) | Mixed (PD: 231 with Ashkenazi Jewish Grandparent, Controls: 97 with Ashkenazi Jewish Grandparent) | DNA sequencing of the full *GBA* gene | Full *GBA* gene | N370S, L444P, 84GG, R496H, IVS2+1, K-27R, E326K, T369M, L461P, V294M, A456P, G241R, Rearrangement exon 8, Q-8H, R44C, N392S, S110A, T410M, F-36V, P387P, E349K | N370S: PD: 36/517 (7.0%), Controls: 4/252 (1.6%)<br>L444P: PD: 7/517 (1.4%), Controls: 1/252 (0.4%)<br>84GG: PD: 4/517 (0.8%), Controls: 0/252 (0%)<br>R496H: PD: 4/517 (0.8%), Controls: 0/252 (0%)<br>IVS2+1: PD: 2/517 (0.4%), Controls: 0/252 (0%)<br>K-27R: PD: 2/517 (0.4%), Controls: 0/252 (0%)<br>E326K: PD: 13/517 (2.5%), Controls: 3/252 (1.2%)<br>T369M: PD: 5/517 (1.0%), Controls: 4/252 (1.6%)<br>L461P: PD: 1/517 (0.2%), Controls: 0/252 (0%)<br>V294M: PD: 1/517 (0.2%), Controls: 0/252 (0%)<br>A456P: PD: 1/517 (0.2%), Controls: 0/252 (0%)<br>G241R: PD: 1/517 (0.2%), Controls: 0/252 (0%)<br>Rearrangement exon 8: PD: 1/517 (0.2%), Controls: 0/252 (0%)<br>Q-8H: PD: 1/517 (0.2%), Controls: 0/252 (0%)<br>R44C: PD: 1/517 (0.2%), Controls: 0/252 (0%)<br>N392S: PD: 1/517 (0.2%), Controls: 0/252 (0%)<br>S110A: PD: 0/517 (0%), Controls: 1/252 (0.4%)<br>T410M: PD: 0/517 (0%), Controls: 1/252 (0.4%)<br>F-36V: PD: 0/517 (0%), Controls: 1/252 (0.4%)<br>P387P: PD: 0/517 (0%), Controls: 1/252 (0.4%)<br>E349K: PD: 0/517 (0%), Controls: 1/252 (0.4%) |
| Clark et al., 2007[84] (17875915) | 278/179 | Mixed (178 PD Jewish, 85 controls Jewish) | DNA sequencing of all *GBA* exons | All *GBA* exons | 84insGG, E326K, T369M, N370S, D409H, R496H, L444P, RecNciI (L444P + A456P + V460V), P175P | All: PD: 38/278 (13.7%), Controls: 8/179 (4.5%)<br>84insGG: PD: 5/278 (1.8%), Controls: 0/179 (0%)<br>E326K: PD: 1/278 (0.4%), Controls: 1/179 (0.6%)<br>T369M: PD: 3/278 (1.1%), Controls: 3/179 (1.7%)<br>N370S: PD: 23/278 (8.3%), Controls: 4/179 (2.2%)<br>D409H: PD: 1/278 (0.4%), Controls: 0/179 (0%)<br>R496H: PD: 1/278 (0.4%), Controls: 0/179 (0%)<br>L444P: PD: 2/278 (0.7%), Controls: 0/179 (0%)<br>RecNciI (L444P + A456P + V460V): PD: 1/278 (0.4%), Controls: 0/179 (0%)<br>P175P: PD: 1/278 (0.4%), Controls: 0/179 (0%) |
| Alcalay et al., 2010[85] (20837857) | 953/0 | Mixed (77 Hispanics, 139 of Jewish ancestry) | DNA sequencing of full *GBA* gene in 90 cases (previously reported); Genotyping for L444P and N370S in 515 | Full *GBA* gene; L444P, N370S | L444P, N370S | L444P: PD: 18/953 (1.9%)<br>N370S: PD: 40/953 (4.2%) |

| | | | | | | |
|---|---|---|---|---|---|---|
| | | | cases; direct sequencing of L444P and N370S in 348 cases | | | |
| Goker-Alpan et al., 2006[86] (16790605) | 28/44 | Mixed | DNA sequencing of full *GBA* gene | All *GBA* exons and flanking introns | N370S | N370S: PD: 1/28 (3.57%), Controls: 0/44 (0%) |
| Sidransky et al., 2009[87] (19846850) | 5691/4898 | Mixed | Variant screening for L444P, N370S or sequencing for all *GBA* exons | L444P, N370S; all *GBA* exons | L444P, N370S, E326K, T369M | Ashkenazi Jews:<br>L444P/N370S: PD (15.3%), Controls (3.4%)<br>Non-Ashkenazi Jews:<br>L444P/N370S: PD (3.2%), Controls (0.6%)<br><br>Center Brazil (N370S, L444P, G377S):<br>PD: 4/65 (6.2%), Controls: 0/264 (0%)<br>Center NYC, USA (Full sequencing):<br>PD: 34/275 (177 AJ) (12.4%), Controls: 3/140. (65 AJ) (2.14%)<br>Center France (N370S, L444P, D409H):<br>PD: 12/297 (4.0%), Controls: 1/251 (0.39%)<br>Center Haifa, IL (D409H, 84GG, V394L, IVS2+1, R496H):<br>PD: 40/162 (162 AJ) (24.7%), Controls: NP<br>Center Italy (L444P, N370S):<br>PD: 11/395 (2.8%), Controls: 1/483 (0.21%)<br>Center Norway (L444P, N370S):<br>PD: 7/311 (2.3%), Controls: 8/473 (1.69%)<br>Center NHGRI, USA (Full sequencing):<br>PD: 29/539 (5.4%), Controls: 6/209 (1 AJ) (2.87%)<br>Center Portugal (Full sequencing):<br>PD: 15/231 (6.5%), Controls: 6/482 (1.24%)<br>Center Rostock, DE (Full Sequencing):<br>PD: 18/298 (6.0%), Controls: 5/212 (2.4%)<br>Center Singapore (L444P, N370S):<br>PD: 8/329 (2.4%), Controls: 0/201 (0%)<br>Center Taiwan (L444P, recNciI, R120W, some full sequencing):<br>PD: 22/559 (3.9%), Controls: 4/377 (1.06%)<br>Center Tel Aviv, IL (84GG, IVS2+1, N370S, V394L, D409H, L444P, R496H, RecTL):<br>PD: 81/420 (419 AJ) (19.3%), Controls: 13/321 (321 AJ) (4.05%)<br>Center Japan (full sequencing):<br>PD: 50/534 (9.4%), Controls: 2/546 (0.37%)<br>Center Tübingen, DE (L444P, N370S):<br>PD: 12/377 (3.2%), Controls: 0/325 (0%)<br>Center Toronto, CA (N370S, K178T, L444P, 84GG, R329C, IVS2+1, recNciiI):<br>PD: 5/88 (2 AJ) (5.7%), Controls: 1/96 (1.0%) |
| Mata et al., 2016[88] (26296077) | 1369/0 | Mixed | DNA sequencing of all *GBA* exons and intron-exons boundaries | All *GBA* exons and intron-exons boundaries | IVS2+1G>A, 84dupG, S125N, T134P, D140H, R163X, N188S, S196P, G202R, F216Y, 914delC, S271G, R359X, N370S, Rec3 (c1263-1317 del, D409H, L444P, A456P, V460V), D409H, L444P, Rec1 (L444P, A456P, V460V), Rec L444P + V460V, V460M, R463C, R496H, R(−32)T, P(−28)S, R44C, G193E, R262H, F316I, G344S, D443N, V460L, S488T, K(−27)R, E326K, T369M | IVS2+1G>A: PD: 2/1369 (0.15%)<br>84dupG: PD: 3/1369 (0.22%)<br>S125N: PD: 1/1369 (0.07%)<br>T134P: PD: 1/1369 (0.07%)<br>D140H: PD: 2/1369 (0.22%)<br>R163X: PD: 1/1369 (0.07%)<br>N188S: PD: 1/1369 (0.07%)<br>S196P: PD: 1/1369 (0.07%)<br>G202R: PD: 1/1369 (0.07%)<br>F216Y: PD: 1/1369 (0.07%)<br>914delC: PD: 1/1369 (0.07%)<br>S271G: PD: 1/1369 (0.07%)<br>R359X: PD: 1/1369 (0.07%) |

| | | | | | | |
|---|---|---|---|---|---|---|
| | | | | | | N370S: PD: 18/1369 (1.31%)<br>Rec3 (c1263-1317 del, D409H, L444P, A456P, V460V): PD: 1/1369 (0.07%)<br>D409H: PD: 1/1369 (0.07%)<br>L444P: PD: 16/1369 (1.17%)<br>Rec1 (L444P, A456P, V460V): PD: 2/1369 (0.22%)<br>Rec L444P + V460V: PD: 1/1369 (0.07%)<br>V460M: PD: 1/1369 (0.07%)<br>R463C: PD: 3/1369 (0.22%)<br>R496H: PD: 2/1369 (0.22%)<br>R(−32)T: PD: 1/1369 (0.07%)<br>P(−28)S: PD: 1/1369 (0.07%)<br>R44C: PD: 1/1369 (0.07%)<br>G193E: PD: 1/1369 (0.07%)<br>R262H: PD: 1/1369 (0.07%)<br>F316I: PD: 1/1369 (0.07%)<br>G344S: PD: 1/1369 (0.07%)<br>D443N: PD: 1/1369 (0.07%)<br>V460L: PD: 1/1369 (0.07%)<br>S488T: PD: 1/1369 (0.07%)<br>K(−27)R: PD: 6/1369 (0.44%)<br>E326K: PD: 69/1369 (5.04%)<br>T369M: PD: 30/1369 (2.19%) |
| Nichols et al., 2009[89] (18987351) | 1325/359 | International | DNA sequencing of all *GBA* exons and corresponding intron/exon boundaries in 96 samples; Variant screening in 1325 cases and 359 controls | All *GBA* exons and intron/exon boundaries | IVS6 589-2A>G, R262H, K303K, E326K, T369M, N370S, L444P, IVS10 1389-3C>G, RecNciI (L444P+A456P+V60V) | IVS6 589-2A>G/R262H/K303K/E326K/T369M/N370S/L444P/IVS10 1389-3C>G/RecNciI (L444P+A456P+V60V): PD: 21/96 (21.9%)<br>All 9 *GBA* variants: PD: 161/1325 (12.2%)<br>All 5 previous *GBA* variants (E326K, T369M, N370S, L444P, RecNcil) : PD: ?/450 (12.6%), Controls: ?/359 (5.3%)<br>E326K: PD: ?/450 (6.2%), Controls: ?/359 (3.1%)<br>T369M: PD: ?/450 (2.3%), Controls: ?/359 (1.1%)<br>N370S: PD: ?/450 (1.4%), Controls: ?/359 (0.8%)<br>L444P: PD: ?/450 (1.9%), Controls: ?/359 (0.0%)<br>A456P/V460V/L444P: PD: ?/450 (0.8%), Controls: ?/359 (0.3%)<br>IVS6 589-2A>G: Controls: 0/359 (0%)<br>R262H: Controls: 0/359 (0%)<br>IVS10 1389- 3C>G: Controls: 0/359 (0%) |
| Liu et al., 2016[90] (27717005) | 2304/0 | International | Depending on the study: Whole exome or targeted sequencing or genotyping of N370S, E326K, T369M | All *GBA* exons; N370S, E326K, T369M | K(-27)R, 84GG, R120W, D140H, G195E, H255Q, R257Q, P266L, R359X, G377S, D409H, L444P, L444R, A456P, N462K, R463C, R463P, N370S, E326K, T369M, E388K | 84GG: PD: 1/1921 (0.05%)<br>R120W: PD: 1/1921 (0.05%)<br>D140H: PD: 8/1921 (0.42%)<br>G195E: PD: 1/1921 (0.05%)<br>H255Q: PD: 1/1921 (0.05%)<br>R257Q: PD: 2/1921 (0.10%)<br>P266L: PD: 1/1921 (0.05%)<br>R359X: PD: 1/1921 (0.05%)<br>G377S: PD: 1/1921 (0.05%)<br>D409H: PD: 1/1921 (0.05%)<br>L444P: PD: 13/1921 (0.68%)<br>L444R: PD: 1/1921 (0.05%)<br>A456P: PD: 1/1921 (0.05%)<br>N462K: PD: 1/1921 (0.05%)<br>R463C: PD: 2/1921 (0.10%)<br>N370S: PD: 28/1921 (1.5%)<br>E326K: PD: 92/1921 (4.79%)<br>T369M: PD: 48/1921 (2.50%)<br>E388K: PD: 2/1921 (0.10%) |
| Stoker et al., 2020[91] (32303560) | 250/0 | NA | DNA sequencing of all *GBA* exons in 250 patients; | All *GBA* exons | N370S, L444P, R463C, G10S, N426K, R48W, R257Q, c.762 18T>A, E326K, T369M, E388K, L119L, c.589 86A>G) | All: PD: 36/250 (14.4%)<br>N370S: PD: 3/250 (1.2%)<br>L444P: PD: 3/250 (1.2%)<br>R463C: PD: 1/250 (0.4%) |

| | | | | | | |
|---|---|---|---|---|---|---|
| | | | genotyping in 127 patients | | | G10S: PD: 1/250 (0.4%)<br>N426K: PD: 1/250 (0.4%)<br>R48W: PD: 1/250 (0.4%)<br>R257Q: PD: 1/250 (0.4%)<br>c.762 18T>A: PD: 8/250 (3.2%)<br>E326K PD: 7/250 (2.8%)<br>T369M: PD: 7/250 (2.8%)<br>E388K: PD: 1/250 (0.4%)<br>L119L: PD: 1/250 (0.4%)<br>c.589 86A>G: PD: 1/250 (0.4%)<br><br>N370S: PD: 4/127 (3.1%)<br>R463C: PD: 1/127 (0.8%)<br>G10S PD: 1/127 (0.8%)<br>T369M PD: 3/127 (2.4%)<br>E326K PD: 2/127 (1.6%)<br>E388K PD: 1/127 (0.8%) |
| Gorostidi et al., 2016[92] (27294386) | 92/0 | NA | Targeted DNA sequencing | NA | K13R, Y244C, T408M | K13R: PD: 1/92 (1.1%)<br>Y244C: PD: 1/92 (1.1%)<br>T408M: PD: 3/92 (3.3%) |
| Mata et al., 2008[93] (18332251) | 721/554 (310 spouses, 244 volunteers) | NA | Genotyping | N370S, L444P | N370S, L444P | N370S: PD: 11/721 (1.5%), Controls: 2/554 (0.4%)<br>L444P: PD: 10/721 (1.4%), Controls: 0/554 (0%) |
| Lwin et al., 2004[94] (14728994) | 57/44 | NA | DNA sequencing of full *GBA* gene | All GBA exons and flanking introns | N370S, L444P, K198T, R329C, T369M, E326K | N370S: PD: 5/57 (8.77%), Controls: 0/44 (0%)<br>L444P: PD: 1/57 (1.75%), Controls: 0/44 (0%)<br>K198T: PD: 1/57 (1.75%), Controls: 0/44 (0%)<br>R329C: PD: 1/57 (1.75%), Controls: 0/44 (0%)<br>T369M PD: 3/57 (5.26%), Controls: 0/44 (0%)<br>E326K PD: 1/57 (1.75%), Controls: 0/44 (0%) |
| Malek et al., 2018[95] (29378790) | 1893/0 | NA | DNA sequencing of all *GBA* exons | All *GBA* exons | L444P, N370S, R463C, G202R, R359S, E326K, T369M, D409H, F213I, G189V, G377S, K157Q, L383Xfs, L66P, M123T, N382Xfs, R163s, R257Q, S173s, E481Xfs, G10S, G325W, R170H, T323I, L175I, L324V, P55S, R262H, R329H, R395C, T267I, L268L, Asp315His, Exon 3 hemizygous deletion, A456P, V460V, D140H, I308T, Ex4 hemizygous deletion | L444P: PD: 30/1893 (1.6%)<br>N370S: PD: 11/1893 (0.6%)<br>R463C: PD: 5/1893 (0.3%)<br>G202R: PD: 2/1893 (0.1%)<br>R359S: PD: 2/1893 (0.1%)<br>E326K: PD: 86/1893 (4.5%)<br>T369M: PD: 35/1893 (1.8%)<br>D409H: PD: 1/1893 (0.05%)<br>F213I: PD: 1/1893 (0.05%)<br>G189V: PD: 1/1893 (0.05%)<br>G377S: PD: 1/1893 (0.05%)<br>K157Q: PD: 1/1893 (0.05%)<br>L383Xfs: PD: 1/1893 (0.05%)<br>L66P: PD: 1/1893 (0.05%)<br>M123T: PD: 1/1893 (0.05%)<br>N382Xfs: PD: 1/1893 (0.05%)<br>R163s: PD: 1/1893 (0.05%)<br>R257Q: PD: 1/1893 (0.05%)<br>S173s: PD: 1/1893 (0.05%)<br>E481Xfs: PD: 1/1893 (0.05%)<br>G10S: PD: 1/1893 (0.05%)<br>G325W: PD: 1/1893 (0.05%)<br>R170H: PD: 1/1893 (0.05%)<br>T323I: PD: 1/1893 (0.05%)<br>L175I: PD: 1/1893 (0.05%)<br>L324V: PD: 1/1893 (0.05%)<br>P55S: PD: 1/1893 (0.05%)<br>R262H: PD: 1/1893 (0.05%)<br>R329H: PD: 1/1893 (0.05%) |

| | | | | | | |
|---|---|---|---|---|---|---|
| | | | | | | R395C: PD: 1/1893 (0.05%)<br>T267I: PD: 1/1893 (0.05%)<br>L268L: PD: 1/1893 (0.05%)<br>Asp315His: PD: 1/1893 (0.05%)<br>Exon 3 hemizygous deletion: PD: 1/1893 (0.05%)<br>A456P: PD: 6/1893 (0.3%)<br>V460V: PD: 6/1893 (0.3%)<br>D140H: PD: 2/1893 (0.1%)<br>I308T: PD: 2/1893 (0.1%)<br>Ex4 hemizygous deletion: PD: 2/1893 (0.1%) |
| Eblan et al., 2005[96] (15716572) | 26/0 | NA | DNA sequencing | NA | D140H, RecNciI | D140H: PD: 1/26 (3.85%)<br>RecNciI: PD: 1/26 (3.85%) |
| Barber et al., 2017[97] (28472425) | 106/283 | NA | Variant screening | N370S, L444P | N370S | N370S: PD: 1/106 (0.9%), Controls: 1/283 (0.4%) |
| Malec-Litwinowicz et al., 2014[98] (25168325) | 138/0 | NA | DNA sequencing of *GBA* exons 8 and 9 | *GBA* exons 8 and 9 | N370S, T369M | N370S: PD: 5/138 (3.6%)<br>T369M: PD: 11/138 (7.9%) |
| McNeill et al., 2012[99] (22577228) | 220/0 | NA | Sanger sequencing of the full *GBA* gene | Full *GBA* gene | N370S, L444P, Recombinant alleles, R496H, V460L, IVS2+1 | N370S: PD: 5/220 (2.27%)<br>L444P: PD: 2/220 (0.91%)<br>Recombinant alleles: PD: 2/220 (0.91%)<br>R496H: PD: 1/220 (0.45%)<br>V460L: PD: 1/220 (0.45%)<br>IVS2+1: PD: 1/220 (0.45%) |
| Graham et al., 2020[100] (31809948) | 229/50 | NA | Nanopore sequencing of full *GBA* gene | Full *GBA* gene | E365K, T408M, D179H, N409S, L335=, R78C | E365K: PD: 12/229 (5.24%), Controls: 0/50 (0%)<br>T408M: PD: 7/229 (3.06%), Controls: 2/50 (4.0%)<br>D179H: PD: 1/229 (0.44%), Controls: 0/50 (0%)<br>N409S: PD: 1/229 (0.44%), Controls: 0/50 (0%)<br>L335=: PD: 1/229 (0.44%), Controls: 0/50 (0%)<br>R78C: PD: 1/229 (0.44%), Controls: 0/50 (0%) |

[*] The reported studies use either the conventional nomenclature for *GBA* alleles excluding the 39-residue signal peptide or refer to the processed protein that includes the 39-residue signal peptide following Human Genome Variation Society (HGVS) recommendation. Articles investigating Norwegian and Scandinavian cohorts are highlighted in gray.
